# Impact of COVID-19 pandemic on diet behaviour among UK adults: a longitudinal analysis of the HEBECO study

**DOI:** 10.1101/2021.10.01.21264008

**Authors:** Samuel J. Dicken, John Joseph Mitchell, Jessica Newberry Le Vay, Emma Beard, Dimitra Kale, Aleksandra Herbec, Lion Shahab

**Author notes:** Correspondence to: Samuel J. Dicken. Joint senior authorship (Aleksandra Herbec, Lion Shahab).

## Abstract

COVID-19 pandemic restrictions impacted dietary habits during the initial months of the pandemic, but long-term effects are unclear. In this longitudinal study, self-selected UK adults (n=1,733) completed three online surveys (May-June, August-September and November-December 2020, with a retrospective pre-pandemic component in the baseline survey), self-reporting sociodemographics, lifestyle and behaviours, including high fat, salt and sugar (HFSS) snacks, HFSS meals and fruit and vegetable (FV) intake. Data were analysed using generalised estimating equations. Monthly HFSS snacks portion intake increased from pre-pandemic levels (48.3) in May-June (57.6, p<0.001), decreased in August-September (43.7, p<0.001), before increasing back to pre-pandemic levels in November-December (49.2, p<0.001). 48.5% self-reported increased (25.9 [95% confidence interval: 24.1, 27.8]) and 47.7% self-reported decreased (24.1 [22.4,26.0]) monthly HFSS snacks portion intakes in November-December compared with pre-pandemic levels. Monthly HFSS meals portion intake decreased from pre-pandemic levels (7.1) in May-June (5.9, p<0.001), being maintained in August-September (5.9, p=0.897), and then increasing again in November-December (6.6, p<0.001), to intakes that remained lower than pre-pandemic levels (p=0.007). 35.2% self-reported increased (4.8 [4.3, 5.3]) and 44.5% self-reported decreased (5.1 [4.6,5.6]) monthly HFSS meals portion intakes in November-December compared with pre-pandemic levels. The proportion meeting FV intake recommendations was stable from pre-pandemic through to August-September (70%), but decreased in November-December 2020 (67%, p=0.034). Increased monthly HFSS snacks intake was associated with female gender, lower quality of life, and - in a time-varying manner - older age and higher HFSS meals intake. Increased monthly HFSS meals intake was associated with female gender, living with adults only and higher HFSS snacks intake. Reduced FV intake was associated with higher body mass index (BMI) and lower physical activity. These results suggest large interindividual variability in dietary change during the first year of the pandemic, with important public health implications in individuals experiencing persistent increases in unhealthy diet choices, associated with BMI, gender, quality of life, living conditions, physical activity and other dietary behaviours.

**What is currently known from previous studies:**

- The introduction of lockdown restrictions impacted on diet behaviours during the initial months of the pandemic.
- Changes in dietary behaviours have been wide ranging, with individuals making healthy and unhealthy dietary changes in high fat, salt and sugar (HFSS) snacks, HFSS meals and fruit and vegetable (FV) intake.
- These dietary changes are associated with multiple sociodemographic, lifestyle and behavioural factors.
- Whether dietary changes persist across longer periods of the pandemic, and which factors are associated with any long-term dietary behaviour change is yet to be identified.

**What this paper adds:**

- Average UK adult intakes of HFSS snacks and meals fluctuated across the pandemic, with the former returning to pre-pandemic levels and the latter remaining below pre-pandemic levels.
- FV intake was stable until the end of 2020, when the proportion meeting recommended intakes declined.
- Across the first year of the pandemic, being female and having a lower quality of life were associated with an increase in HFSS snacks intake, whereas the association of age and HFSS meals intake with HFSS snacks intake varied across the pandemic. Living with adults only and having a higher intake of HFSS snacks were associated with an increase in HFSS meals intake. A higher BMI and lower physical activity level were associated with reduced FV intake.

## 1. Introduction

2020 has seen widespread disruption to the lives of individuals across the globe due to the COVID-19 pandemic. In the UK, pandemic restrictions were imposed from late March 2020 onwards. Since then, varying levels of restrictions have impacted how individuals and societies live their lives [1]. Restrictions were slowly eased from June until September, before progressively becoming stricter, with full lockdown conditions in December 2020. Diet is a major factor influencing bodyweight, blood pressure, metabolic health and the risk of non-communicable disease. As such, diet is one of the largest contributors to the burden of disease (as measured using disability-adjusted life-years (DALYs)) globally and in the UK [2,3]. The widespread disruption to people’s lifestyles from the COVID-19 pandemic may result in significant shifts in health behaviours, including diet.

Factors that can impact on energy intake and dietary behaviour including food accessibility, changes in work life, home life, stress and other health behaviours including sleep, physical activity, smoking and alcohol consumption have been affected by COVID-19 restrictions [4–7]. Closures of restaurants and fast food outlets, increased usage of food delivery services, changes in the affordability and availability of foods alongside disruptions to the home and working environment, as well as changes in employment status may all impact on diet behaviour during the pandemic [8– 13]. Indeed, initial reports find that a significant proportion of adults have altered their food choices and dietary habits at the start of the pandemic compared to pre-pandemic food choices and habits [9,14–16]. Despite no overall change in diet quality, there has been large interindividual variability [1,5,14,17,18]. A scoping review of 23 studies (17 cross-sectional) from the initial months of the pandemic demonstrate that individuals are making favourable and unfavourable changes to their diet, including changes in snacking, high fat, salt or sugar (HFSS) food intake, and fruit and vegetable (FV) intake [14]. Significant proportions of people have increased their overall food intake and are snacking more [5,7,14,18,19], but equally people have also decreased their overall food intake and are snacking less [1,9,18,20].

These changes in diet behaviours during the pandemic are associated with several factors including age, gender, body mass index (BMI), physical activity and experiencing a larger psychological impact and larger shifts in lifestyle as a result of lockdown restrictions [4,14,21–27]. Given the relationship between a sub-optimal diet and relative risk of cardiovascular disease, cancer and all-cause mortality, there could be serious long-term public health consequences if the initial unfavourable changes in dietary behaviours during the pandemic are maintained [28,29]. Understanding the dietary changes that have occurred, and the key predictors associated with these changes is important to identify at-risk groups of unhealthful dietary change, to inform future interventions, develop targeted approaches and guide efficient resource allocation. Given the impact of culture on diet and the specific impacts of lockdown restrictions across the globe, it is important to assess the longitudinal impact, including in the UK. However, studies to date assessing the influence of COVID-19 lockdown restrictions on dietary behaviours in UK adults have largely been cross-sectional, undertaken during the initial months of lockdown [7,14,18]. Some cross-sectional analyses show dietary changes during August-October 2020 compared to the pre-pandemic period [1,30,31], and changes in consumer habits during November-December 2020 [10]. One longitudinal UK study analysing food purchases found increased calorie intake across 2020 [16], but longitudinal analyses are scarce [16,32]. Dietary changes compared to pre-pandemic levels and key predictors of any change during the first year of the pandemic to December 2020 in UK adults are largely unknown.

The HEalth BEhaviours during the COVID-19 pandemic (HEBECO) study is a longitudinal UK cohort assessing the impact of the COVID-19 pandemic on health behaviours, and their influences. The objective of this study was to address the following research questions:

RQ1. What was the average (i) HFSS snacks, (ii) HFSS meals and (iii) FV intake in UK adults before, at the beginning of, and at 3-months and 6-months follow-up during the COVID-19 pandemic?

RQ2. To what extent are sociodemographic, COVID-19-related and behavioural factors associated with a change in (i) HFSS snacks, (ii) HFSS meals intake or (iii) a reduction in FV intake across 6-months of follow-up during the COVID-19 pandemic compared with pre-pandemic intakes in UK adults?

## 2. Materials and Methods

### 2.1. Study design

The study design has been previously reported [33]. Briefly, this study is a longitudinal analysis of data from an online study of adults, the HEBECO study (https://osf.io/sbgru/). The study was approved by the Ethics Committee at the UCL Division of Psychology and Language Sciences (CEHP/2020/759). Participants were self-selected and gave consent prior to data collection. The full recruitment strategy is available online (https://osf.io/sbgru/). Data were captured and managed by the REDCap electronic data system at UCL [34,35]. The surveys used in this analysis cover a period of 8 months since the beginning of the pandemic (May to December 2020), as well as a retrospective survey at baseline of the pre-pandemic period. Baseline data was collected between 5^th^ May and 14^th^ June 2020 (inclusive). The 3-months follow-up survey corresponds to the periods of eased pandemic restrictions in the UK during August-September 2020, and the 6-months follow-up survey corresponds to the tighter restrictions in the UK during November-December 2020. The study protocol and statistical analysis plan were pre-registered on the Open Science Framework prior to analysis (https://osf.io/279zd/). Deviations from the pre-registered protocol are described in the Supplementary Materials. The main study protocol (https://osf.io/mav3y/) provides further detail on the survey.

### 2.2. Study sample

The analysis uses data from UK adults (18+) who completed baseline data collection and provided data of interest at the 6-months follow-up survey as a minimum, for the outcome variables defined below.

### 2.3. Measures

Full details of outcome and predictor measures can be found in the Supplementary Materials, and have been previously defined [33].

#### 2.3.1 Outcomes

Participants were asked at baseline, ‘*Before COVID-19, how often did you usually eat or drink*…’ for nine food items. Eight HFSS food items ((i) ready meals, (ii) fast food, (iii) takeaways, (iv) sugary or sweetened drinks, (v) sweets or chocolate, (vi) cakes and biscuits, (vii) desserts and (viii) savoury snacks), and one item for fruit and vegetable intake. For each food item, respondents could answer on a 7-point scale: ‘A few times per day’, ‘Once a day’, ‘A few times per week’, ‘Once a week’, ‘A few times per month’, ‘Once a month’, ‘Less often/never’ and ‘Not sure’. Participants were also asked at baseline ‘*Since COVID-19, how often did you usually eat or drink*…’, and then at 3-months and 6-months follow-up, *‘In the past month, how often did you usually eat or drink*…’ for the same nine food items. The HEBECO study food item questions are based on previous research study survey questions, and derived from Public Health England’s sugar reduction programme definitions as policy relevant measures [36–38].

To estimate monthly portion intake frequency, responses for all food item questions were converted into monthly portion frequencies, based on previous research [33]. Assuming a minimum of four weeks per calendar month, an answer of ‘A few times per day’ was scaled up to 56 portions per month (i.e. two daily portions x seven days x four weeks), ‘Once a day’ was scaled up to 28 portions per month, ‘Few times per week’ as 12 portions per month, ‘Once a week’ as four portions per month, ‘Few times per month’ as two portions per month, ‘Once a month’ as one portion per month and ‘Less often/never’ as 0.5 portions per month.

Using the above monthly portion intake frequencies, (iv) sugary or sweetened drinks, (v) sweets or chocolate, (vi) cakes and biscuits, (vii) desserts and (viii) savoury snacks monthly frequencies were summed to produce a ‘HFSS snacks intake’ monthly portion frequency. (i) Ready meals, (ii) fast food and (iii) takeaways monthly frequencies were summed to produce a ‘HFSS meals intake’ monthly portion frequency. The change scores ‘Change in self-reported HFSS snacks intake’ and ‘Change in self-reported HFSS meals intake’ used as outcomes in RQ2 were computed from pre-pandemic HFSS snacks and HFSS meals intakes retrospectively reported at baseline, which were deducted from HFSS snacks and HFSS meals intakes reported at the time of the baseline survey, and at 3- and 6-months follow-up surveys.

FV intake was converted into a binary outcome variable, grouped into ‘Consuming a few portions per day of fruit and vegetables’ (responses of ‘A few times per day’) vs ‘Less than a few portions per day’ (all other responses besides ‘A few times per day’). This cut-off was used to reflect health recommendations for several daily portions of fruit and vegetables [39]. The binary change score used as the outcome in RQ2 was computed as a categorical reduction (‘Reduced intake’ vs ‘All other’) in FV intake at the time of the baseline survey, and at 3- and 6-months follow-up surveys, compared to pre-pandemic levels retrospectively reported at baseline.

HFSS snacks intake was the primary outcome of interest, given that a systematic review identified large changes in snacking during the initial months of the pandemic [18], which in turn have been associated with weight change during the pandemic [5,40,41].

#### 2.3.2. Explanatory variables

##### 2.3.2.1. Time-invariant

Explanatory variables recorded at baseline included **gender** (female vs all other), **age** (continuous), **ethnicity** (white vs all other), **occupation and work from home** (categorical: unemployed (which includes retired persons and full-time parents/carers), employed and working from home, employed and not working from home), **living arrangements** (living alone, living with children (with or without adults), living with adults only) and a **socioeconomic score**. The socioeconomic score (categorical score from 0-3) was based on household income, housing status and level of education; participants scored 0 if they had an income <£50,000, lived in unowned housing and had no higher education, or scored 1, 2 or 3 if participants met 1, 2 or all 3 of having an income of ≥£50,000, owning their housing/having a mortgage, or having higher education.

Sensitivity analyses also included a **composite eating behaviour score**. This measure was not included in the main RQ2 analysis as not all participants were shown the question to reduce participant burden. At baseline, participants were asked to what extent they agreed with the following statements: *‘I eat unhealthy food out of boredom’*, ‘*I eat unhealthy food because I’m stressed’*, ‘*I eat unhealthy food because it’s comforting’* on a 0-100 scale, where 0 = completely disagree; 50 = neutral; and 100 = completely agree. A continuous mean score (0-100) was computed for eating for comfort, stress and from boredom. A higher score indicated eating unhealthy food out of boredom, stress or comfort.

##### 2.3.2.2. Time-variant

Explanatory variables reported at baseline, 3- and 6-months follow-up surveys included **BMI** (continuous: weight in kilograms divided by height in metres squared), **isolation status** (total/some isolation vs general/no isolation), **quality of life**, an average continuous rating from 1-5 of quality of living, well-being, social and family relationships (1 = poor, 5 = excellent), and health behaviours as detailed below.

**Physical activity** was a continuous measure of MET-hours per week. At each time point, participants self-reported the number of days they performed strengthening physical activity (SPA) per week, and the number of days and average duration of a session of moderate or vigorous physical activity (MVPA) per week. The number of days performing SPA per week was multiplied by an average session duration of 45 minutes and multiplied by 4 to convert to MET-minutes per week, to reflect the nature of SPA as moderate-to high-intensity bouts, interspersed with rest periods [42]. The 45-minute length reflects the American College of Sports Medicine recommendations for the typical number of exercises, reps, sets and duration of rest periods for a resistance training session [43,44]. MVPA number of days per week was multiplied by the self-reported average session length, and then multiplied by 6 to convert to MET-minutes per week, as an average of moderate and vigorous physical activity [45]. Scores were then summed for SPA and MVPA and divided by 60 to produce a MET-hours per week score. An upper limit of 4 standard deviations above the mean MET-hours per week was applied, as some individuals self-reported activity levels not physically possible. This upper limit corresponded to ∼8 hours of moderate physical activity per day, which is several standard deviations above the physical activity levels reported from large observational studies [46].

**Alcohol consumption** was based on government low-risk drinking recommendations (≤14 weekly alcohol units vs >14 weekly alcohol units) [47] and **smoking status** was based on the self-reported use of tobacco or cigarettes (yes vs no).

**HFSS snacks intake, HFSS meals intake** and **FV intake** were also used as continuous explanatory variables (but excluded in analyses of the same kind, e.g. HFSS snacks intake was excluded from ‘Change in self-reported HFSS snacks intake’ analyses).

### 2.4. Statistical Analysis

Statistical analysis was conducted in SPSS Statistics version 27 (IBM). Significance was defined as p<0.05.

#### 2.4.1. RQ1

We described baseline participant characteristics in RQ1 (weighted participant characteristics, based on census data from the Office for National Statistics for age, gender, country of living, ethnicity, education and income are presented in the Supplementary Materials [48]).

We reported the unweighted means with 95% confidence intervals (95%CI) for HFSS snacks and HFSS meals monthly portion intake and the proportion consuming a few portions of FV per day at each timepoint. We also reported the percentage of the sample increasing or decreasing HFSS snacks or HFSS meals intake, or categorically increasing or decreasing FV intake from the reference timepoint (pre-pandemic levels reported at baseline, levels during May-June 2020 reported at baseline, or at 3-months follow-up). Lastly, we reported the mean change in portion frequency (with 95%CI) in those increasing or decreasing HFSS snacks or HFSS meals intake between timepoints.

Given that more distantly spaced participant measures over time are expected to be less closely correlated [49], an unadjusted, unweighted generalised estimating equation (GEE) using the AR(1) covariance structure was used to assess changes in self-reported HFSS snacks and HFSS meals monthly portion intake over time, using pairwise time comparisons between timepoints, adjusted for with sequential Šidák correction.

#### 2.4.2. RQ2

GEE models were used to determine the association between the explanatory variables and (i) changes in HFSS snacks intake and (ii) changes in HFSS meals intake across the follow-up period. The GEE models for a change in HFSS snacks and HFSS meals monthly portion intake used the identity link function for a linear scale response, as the change scores were normally distributed continuous outcome variables. The GEE models for FV used a binary logistic model and logit link function for the binary outcome variable (reduced FV intake vs all other).

Univariate GEE models were computed to determine the association between each explanatory variable and changes in HFSS snacks intake, changes in HFSS meals intake, and a reduction in FV intake. Each explanatory variable model was adjusted for a main effect of time and for an ‘explanatory variable*time’ interaction. Fully adjusted GEE models containing all explanatory variables were then computed.

All significant explanatory variable*time interactions were then added to the fully adjusted GEE model containing all explanatory variables, to assess temporal differences in the association of explanatory variables with continuous changes in HFSS snacks and HFSS meals monthly portion intake, or a reduction in FV intake over time. The time variable was categorical, as the trajectory of change in dietary intakes was not expected to be linear [50]. Explanatory variable*time interactions were retained in the full GEE model if they improved goodness of fit (Quasi-likelihood under Independence Model Criterion (QIC) QIC >2) over the full GEE model without interactions, and the interaction itself remained significant (p<0.05).

Independent variables were retained after checking for collinearity using Pearson correlations, with all correlations *r*<0.4.

For binary outcomes using the logit link function, linearity of logit assumptions were checked, as detailed in the Supplementary Materials.

#### 2.4.3. Sensitivity analyses

The supplementary analyses used complete cases only for all dietary measures at baseline, 3-months and 6-months follow-up. Further analyses were then conducted for the primary outcome of HFSS snacks intake using two binary logistic GEE models with logit link function for an ‘increase’ in HFSS snacks intake vs ‘all other’, and a ‘decrease’ in HFSS snacks intake vs ‘all other’. A change was defined as an increase or decrease in intake by 10% or more from pre-pandemic intakes retrospectively self-reported at baseline, as a cut-off used in a previous large study of dietary intakes during the pandemic [4].

Analyses were repeated for the sub-sample of participants self-reporting eating behaviour measures. Univariate GEE models were computed for changes in HFSS snacks intake, changes in HFSS meals intake and a reduction in FV intake outcomes using the eating behaviour score. The eating behaviour score was also added to the fully adjusted GEE models (with and without significant time interactions from the univariate models).

Multiple observations in the literature have indicated that gender, smoking and physical activity are likely related to changes in snacking behaviour during the pandemic [4,7]. Bayes Factor analyses were pre-registered online in the event of non-significant findings for gender, smoking and physical activity with change in HFSS snacks intake in the main analysis (https://osf.io/279zd/). Snacking Bayes Factors prior mean differences were obtained from differences in change in daily energy intake from before COVID-19 to since COVID-19 between predictors and converted into snacking portions per month (further details can be found in the Supplementary Materials). Alternative hypotheses were modelled using a half-normal distribution with a peak at zero, given that smaller effect sizes nearer to the null are more likely than larger effect sizes [51,52]. The standard deviation (SD) was set to 5.32 for gender, 7.35 for physical activity and 11.17 for smoking for a change in HFSS snack portions per month [4]. Bayes factors were calculated using an online calculator: http://bayesfactor.info/.

## 3. Results

Out of a total of 2,992 UK adult participants recruited into the HEBECO baseline survey, 1,733 (weighted = 1,532) participants met the inclusion criteria for analyses. The unweighted baseline characteristics are shown in table 1 (for weighted characteristics, see Supplementary table S1). Included participants were more likely to be female, older, of white ethnicity, have a higher BMI, be unemployed (which includes retired persons and full-time parents/carers), have a higher socioeconomic score, live with adults only, be in total or some isolation, have a higher quality of life score, consume fewer HFSS meals, consume more FV and be less likely to smoke.

**Table 1.** Unweighted baseline participant characteristics for included, excluded and total samples.

|  | Total Sample | Analytical Sample | Excluded sample |  |
| --- | --- | --- | --- | --- |
|  | % | % | % | p-value |
| **N** | 2992 | 1733 | 1259 |  |
| Gender |  |  |  | **0.001** |
| All other | 31.4% | 28.9% | 34.8% |  |
| Female | 68.6% | 71.1% | 65.2% |  |
| Ethnicity |  |  |  | **<0.001** |
| All other | 6.3% | 4.4% | 8.8% |  |
| White | 93.7% | 95.6% | 91.2% |  |
| Mean BMI [SD] N=2783 | 26.1 [5.2] | 26.3 [5.2] | 25.7 [5.1] | **0.003** |
| Mean Age [SD] | 47.9 [15.5] | 51.5 [14.3] | 43.0 [15.6] | **<0.001** |
| Occupation and work from home N=2990 |  |  |  | **<0.001** |
| Unemployed (including retired persons and full-time parents/carers) | 28.6% | 32.6% | 23.1% |  |
| Employed working from home | 51.6% | 49.1% | 55.1% |  |
| Employed not working from home | 19.8% | 18.3% | 21.9% |  |
| Socioeconomic score |  |  |  | **<0.001** |
| Income <£50k, unowned Housing and no higher education | 4.9% | 3.7% | 6.5% |  |
| 1 of: ≥£50K income, housing ownership/mortgage or higher education | 27.8% | 24.8% | 31.9% |  |
| 2 of: ≥£50K income, housing ownership/mortgage or higher education | 38.5% | 41.1% | 34.9% |  |
| All of: ≥£50K income, housing ownership/mortgage and higher education | 28.8% | 30.5% | 26.6% |  |
| Living conditions |  |  |  | **<0.001** |
| Alone | 16.8% | 17.2% | 16.4% |  |
| With children (with or without adults) | 19.5% | 16.7% | 23.4% |  |
| With adults only | 63.6% | 66.1% | 60.2% |  |
| Isolation status N=2946 |  |  |  | **0.010** |
| Total or some isolation | 79.3% | 80.9% | 77.0% |  |
| General or no isolation | 20.7% | 19.1% | 23.0% |  |
| Mean Quality of Life [SD] | 3.4 [0.8] | 3.4 [0.8] | 3.3 [0.8] | **<0.001** |
| Mean HFSS Snacks portions per month N=2609 | 58.4 [45.2] | 57.6 [44.4] | 60.0 [46.8] | 0.214 |
| Mean HFSS Meals portions per month N=2618 | 6.6 [8.3] | 5.9 [6.7] | 8.1 [10.6] | **<0.001** |
| Mean Fruit and vegetables portions per month N=2647 | 44.0 [18.0] | 45.3 [17.2] | 41.5 [19.3] | **<0.001** |
| Mean physical activity MET-Hours per week N=2804 | 20.6 [20.8] | 21.4 [21.0] | 19.2 [20.3] | **0.008** |
| Alcohol consumption N=2772 |  |  |  | 0.290 |
| ≤14 weekly units | 81.0% | 81.6% | 79.9% |  |
| >14 weekly units | 19.0% | 18.4% | 20.1% |  |
| Smoking status |  |  |  | **<0.001** |
| Yes | 18.6% | 13.0% | 26.3% |  |
| No | 81.4% | 87.0% | 73.7% |  |

*\*P-values are for comparisons between the analytical and excluded samples. 90 participants were excluded due to reporting 'not sure to the diet questions at baseline or 6-months follow-up. SD=Standard deviation.*

### 3.1. What were the intakes of HFSS snacks, meals and FV in UK adults before, at the beginning of, and at 3-months and 6-months follow-up during the COVID-19 pandemic?

HFSS snacks monthly portion intake increased from before the pandemic to the start of the pandemic (by May-June 2020) (48.3 to 57.6 per month, p<0.001) (Figure 1). At 3-months follow-up (by August-September 2020), HFSS snacks monthly portion intake significantly decreased (to 43.7 per month, p<0.001). At 6-months follow-up (by November-December 2020), HFSS snacks monthly portion intake significantly increased (to 49.2 per month, p<0.001), to intakes that were not significantly different to retrospectively reported intakes before the pandemic (p=0.297).

**Figure 1.**
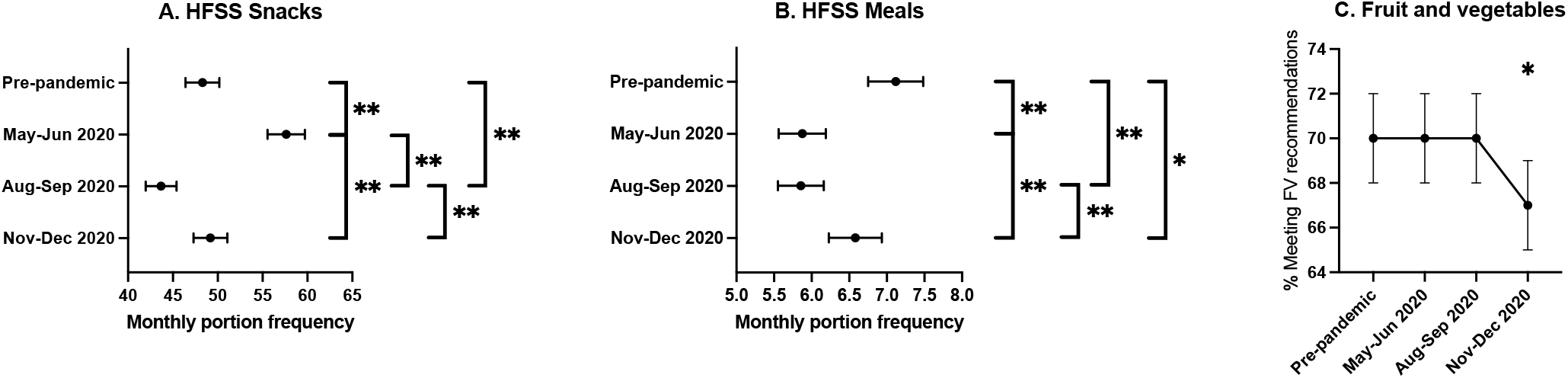
Means, 95% confidence intervals and pairwise comparisons between pre-pandemic, baseline (May-June 2020), 3-month (August-September 2020) and 6-month (November-December 2020) follow-up surveys for (A) HFSS snacks, (B) HFSS meals and (C) FV intake. *\*\*Pairwise comparisons between timepoints were significant at the 0.005 level. *Pairwise comparisons between timepoints were significant at the 0.05 level.*

HFSS meals monthly portion intake decreased from before the pandemic to the start of the pandemic (by May-June 2020) (7.1 to 5.9 per month, p<0.001) (Figure 1). At 3-months follow-up (by August-September 2020), HFSS meals monthly portion intake was maintained (5.9 per month, p=0.897). At 6-months follow-up (by November-December 2020), HFSS meals monthly portion intake significantly increased (5.9 to 6.6 per month, p<0.001), to intakes that were significantly lower than retrospectively reported intakes before the pandemic (7.1 to 6.6 per month, p=0.007).

For FV intake, 70% [95%CI: 68,72] were consuming a few portions per day before the pandemic, at the start of the pandemic [95%CI: 68,72] (May-June 2020) and at 3-months follow-up [95%CI: 68,72] (August-September 2020), but significantly decreased to 67% [95%CI: 65,69] (p=0.034) at 6-months follow-up (November-December 2020) (Figure 1).

From the pre-pandemic period to 6-months follow-up (November-December 2020), 48.5% of individuals self-reported an increase in HFSS snacks intake, by an average of 25.9 [95%CI: 24.1,27.8] portions per month. A similar proportion (47.7%) reported a decrease in HFSS snacks intake, by an average of 24.1 [95%CI: 22.4,25.9] portions per month (Table 2). From pre-pandemic to 6-months follow-up, 35.2% of individuals self-reported an increase in HFSS meals intake, by an average of 4.8 [95%CI: 4.3,5.3] portions per month. 44.5% self-reported a decrease in HFSS meals intake, by an average of 5.1 [95%CI: 4.6,5.6] portions per month. For FV intake, 11.4% were no longer meeting daily FV intake recommendations, and 8.4% were now meeting daily FV intake recommendations at 6-months follow-up compared with pre-pandemic levels.

**Table 2.**
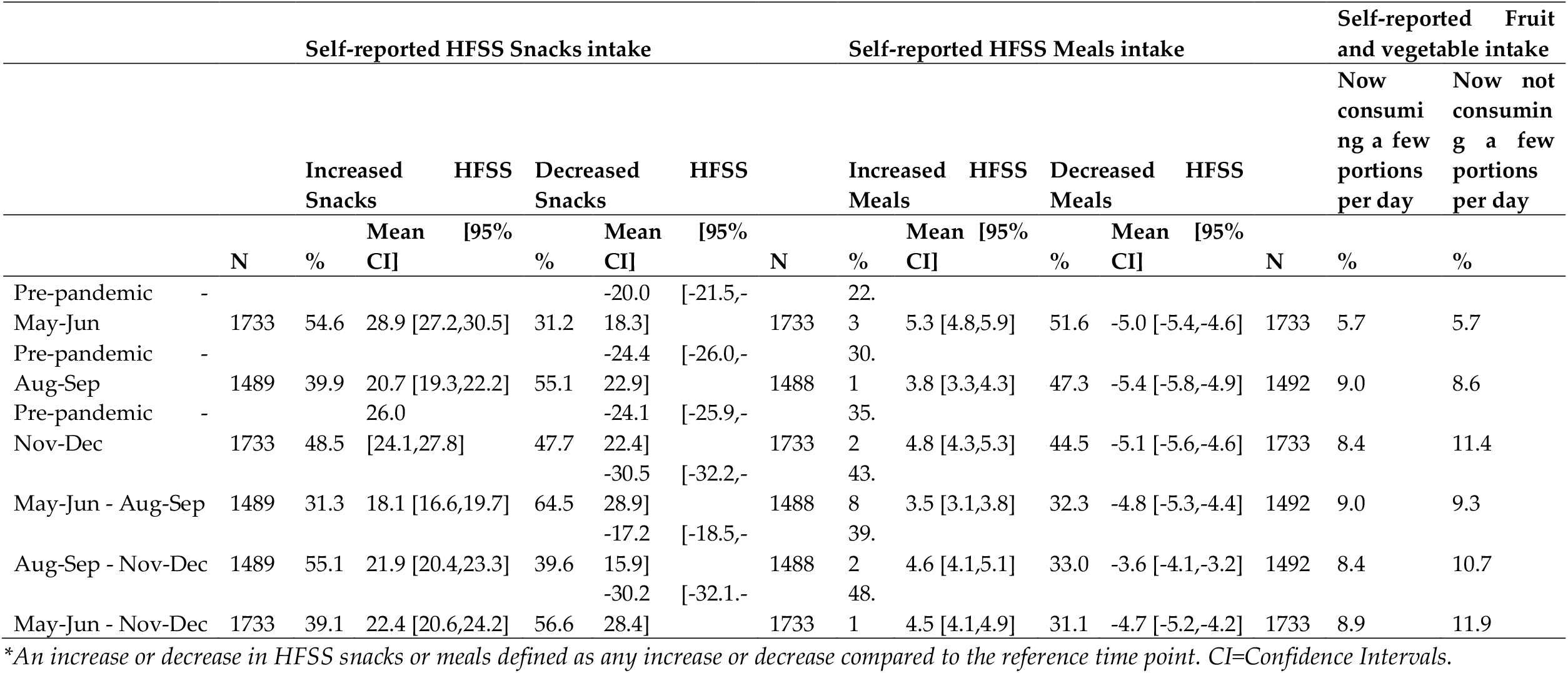
Unweighted proportions increasing or decreasing HFSS snacks, HFSS meals and fruit and vegetables intake and mean changes with 95% confidence intervals in HFSS snacks and HFSS meals intake.

The changes in portion frequency consumption across the pandemic for each food item is shown in Supplementary figure S1.

### 3.2. Which explanatory variables were associated with changes in monthly HFSS snacks and meals intakes, or a reduction in FV intake in UK adults across 6-months of follow-up during the COVID-19 pandemic?

In the unadjusted GEE models (Supplementary table S2), female gender, higher baseline BMI, total or some isolation, a lower quality of life score, a higher HFSS meals intake and lower physical activity levels were significantly associated with an increase in self-reported HFSS snacks intake across 6-months of follow-up during the pandemic. In the fully adjusted GEE model (Table 3), female gender (B=6.568 [95%CI: 3.653,9.483]) and a lower quality of life score (B=-2.882 [95%CI: - 4.387,-1.377]) were associated with an increase in monthly HFSS snacks intake across the pandemic. HFSS meals intake was also significantly associated with a change in monthly HFSS snacks intake across the pandemic, but this was time-varying. Similarly, age showed significant time interactions and improved model fit.

**Table 3.**
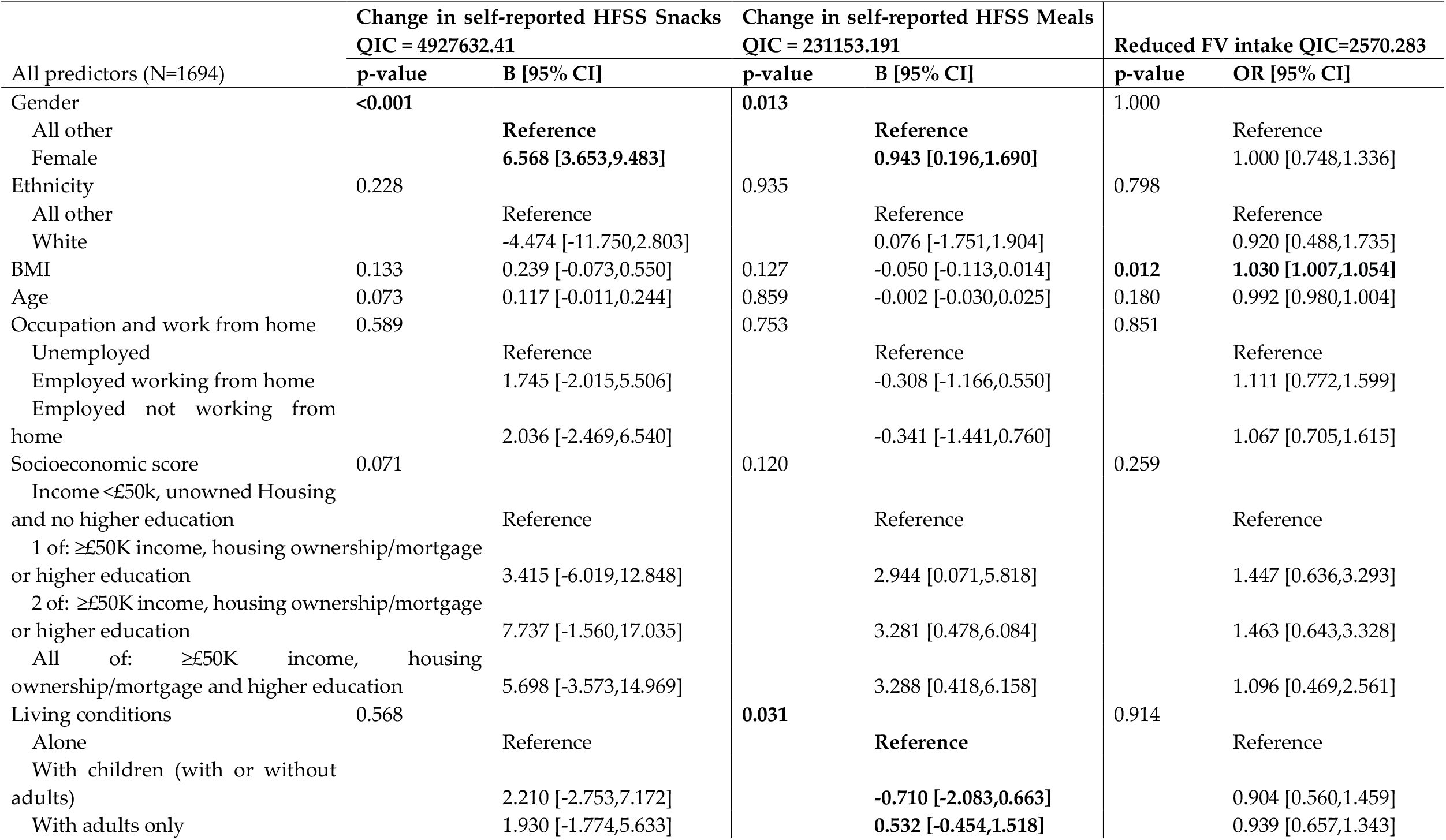
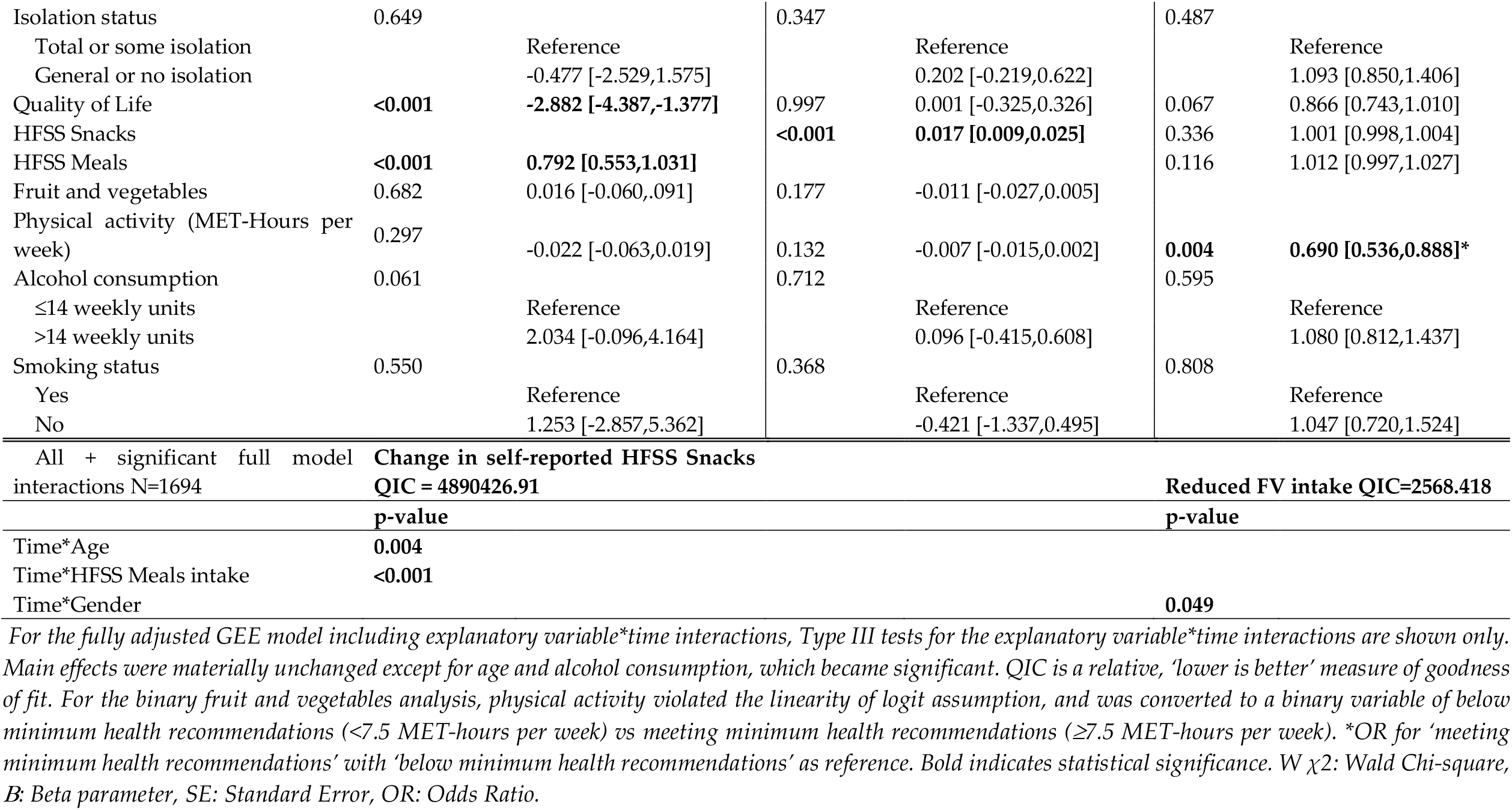
Fully adjusted GEE model containing all predictor variables and the fully adjusted GEE model including significant explanatory variable*time interactions.

Figure 2 exhibits the time-varying associations of age and HFSS meals intake with a change in HFSS snacks intake across the pandemic. All ages increased HFSS snacks intake at the start of the pandemic, but younger ages tended to decrease HFSS snacks intake during the latter months of 2020 (November-December 2020), whereas older ages tended to maintain or increase intakes from pre-pandemic levels. Higher HFSS meals intakes were associated with larger increases in HFSS snacks intake at the start of the pandemic (May-June), with higher and lower HFSS meals intakes decreasing HFSS snacks intake at 3-months follow-up (August-September). Individuals with a higher HFSS meals intake then returned to, or increased HFSS snacks intake above pre-pandemic levels at 6-months follow-up, whereas those with lower HFSS meals intakes maintained the reduced HFSS snacks intake.

**Figure 2.**
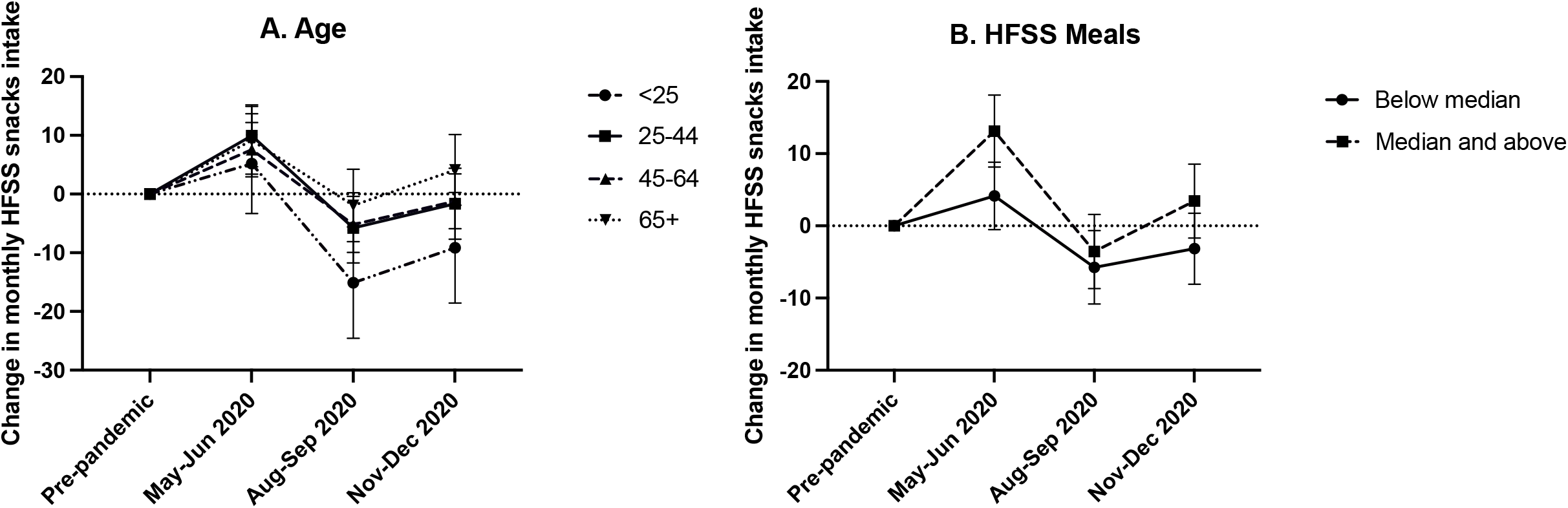
Graphical illustrations of the time-varying associations of (A) age and (B) HFSS meals intake with change in monthly HFSS snacks intake at baseline (May-June 2020), 3-months (August-September 2020) and 6-months follow-up (November-December 2020) compared with pre-pandemic intakes. *Age categories: <25, 5.2%; 25-44, 24.1%; 45-64, 52.2%; >65, 18.6%. Median monthly HFSS meals intake at baseline = 3.5, at 3-months follow-up = 3.5, at 6-*

In the unadjusted GEE models of HFSS meals intake (Supplementary table S2), female gender and a higher HFSS snacks intake were associated with an increase in HFSS meals across the pandemic. In the fully adjusted GEE model (Table 3), female gender (B=0.943 [95%CI: 0.196,1.690]), living with adults only (B=0.532 [95%CI: -0.454,1.518]) and higher intakes of HFSS snacks (B=0.017 [95%CI: 0.009,0.025]) were associated with an increase in HFSS meals intake across the pandemic. There were no significant explanatory variable*time interactions.

In the unadjusted GEE models of FV intake (Supplementary table S2), higher BMI, lower socioeconomic score, general or no isolation (compared with total or some isolation), lower quality of life, higher HFSS meals intake and lower physical activity levels were associated with a reduction in FV intake. In the fully adjusted GEE model (Table 3), reduced FV intake across the pandemic was associated with a higher BMI (odds ratio (OR)= 1.030 [95%CI: 1.007,1.054]) and lower physical activity levels (OR= 0.690 [95%CI: 0.536,0.888]). A gender interaction with time while significant, did not improve model fit and was not retained.

### Sensitivity Analyses

Complete case analyses of RQ1 and RQ2 demonstrated materially unchanged differences in HFSS snacks and HFSS meals intakes across timepoints and in adjusted analyses compared with the main analysis (Supplementary tables S3, 4 and 5). For a change in FV intake, however, the gender interaction with time improved model fit and remained significant when added to the fully adjusted model, as shown in Supplementary figure S2.

Unadjusted analyses of binary outcomes for HFSS snacks used in the sensitivity analysis are presented in Supplementary table S6. In the full binary logistic GEE models for an increase or decrease in HFSS snacks intakes vs all other, female gender, a higher BMI, a lower quality of life score and a higher HFSS meals intake were more likely to increase HFSS snacks intake across the pandemic (Supplementary table S7). Non-female gender, younger age, higher quality of life, lower HFSS meals intake and low-risk alcohol consumption were associated with decreased HFSS snacks intake across the pandemic. Age and alcohol consumption interactions had a time-varying impact on increased HFSS snacks intake vs all other (Supplementary figure S3). There were no significant explanatory variable*time interactions for a decrease in HFSS snacks vs all other (Supplementary table S6).

Unadjusted analyses of the eating behaviour score are also presented in Supplementary table S8. In fully adjusted GEE models including the eating behaviour score, a higher eating behaviour score was significantly associated with an increase in HFSS snacks intake (Supplementary table S9). The eating behaviour score had a time-varying impact (Supplementary figure S4).

Because of the null findings reported here for smoking status on a change in HFSS snacks intake, Bayes Factors were calculated. The Bayes Factor suggests the data provided evidence for no effect of smoking status (BF=0.32) on change in HFSS snacks intake.

## 4. Discussion

In this study of a sample of UK adults, initial average trends in HFSS snacking and meal consumption are not maintained across the pandemic. There is substantial interindividual variability, with some individuals experiencing long-term, unhealthy dietary behaviour changes. Combined, our results suggest that gender, BMI, living conditions, quality of life, other dietary behaviours and physical activity are associated with adverse dietary behaviour changes.

### In context of COVID-19 research

Reviews from the initial months of the pandemic indicate an overall trend for increased food consumption [7,53], with increased FV consumption in some individuals, but also increased HFSS foods consumption in others [6,54]. In particular, increased snacking has been identified during the start of the pandemic [18,54–56], with a greater proportion of individuals increasing than decreasing HFSS snacks intake [18,54], as with our results.

Dietary changes can have both protective and harmful impacts on health and wellbeing. Understanding the contexts associated with dietary changes is central to developing targeted interventions. Female gender, higher BMI, lower quality of life, reduced physical activity and experiencing a greater negative impact from the pandemic have been previously recognised as predictors of unhealthy dietary changes during the pandemic [55,57,58]. Our study builds upon the current literature, showing these factors are important long-term predictors of adverse dietary change during the pandemic.

Pandemic-related dietary changes may have occurred from shop closures, or changes in food availability, accessibility or shopping habits [31,55]. Increased HFSS snacking at the start of the pandemic may have resulted from convenience, or greater inconvenience of accessing fresh foods [55,59]. Given their high availability, affordability and long shelf-life, individuals may have prioritised HFSS snacks over fresh foods from the uncertainty over food supply during the pandemic [55,59–61].

In our study, a lower quality of life or eating through stress, boredom or comfort (eating behaviour score) was associated with unhealthy changes in HFSS foods intake during the first year of the pandemic. Individuals tend to consume more palatable and less healthy foods during stressful life periods [62]. A greater decline in mental health or increased stress, boredom or anxiety from COVID-19 has been associated with increased ultra-processed, HFSS foods intake, decreased FV intake and using snacking as a coping mechanism [9,14,32,55,56,63–65]. The increased HFSS snacking at the start of the pandemic may reflect such maladaptive coping mechanisms [63,66]. For some individuals, the increase may have been maintained through strengthening of a cue-trigger-reward feedback cycle and habit formation [67].

We found that living with adults was associated with increased HFSS meals intake. Higher numbers of adults were shown to consume more meals per day during the second COVID-19 wave in the UK (October 2020) compared with pre-pandemic, with increases in both ready meals and homemade meals [30]. More shared mealtimes during lockdown may have altered eating behaviours [68], with existing or changing HFSS meal habits of some adults in the household potentially influencing the dietary habits of others [68].

### Policy implications

The World Health Organisation recommends limiting HFSS foods to reduce the risk of weight gain, cardiovascular disease and high blood pressure [55,69]. In the UK, average free sugar and saturated fat consumption exceeds recommendations, and average FV intakes are below recommendations [31,70]. This study suggests the pandemic is associated with long-term adverse changes in dietary behaviours, which could amplify the existing sub-optimal dietary patterns of UK adults. A poor diet is the largest behavioural risk factor for DALYs lost [3] and second only to smoking for years of life lost [71], indicating a strong need for policy action to help individuals make healthy dietary choices. The pandemic has impacted people differently, therefore strategies need to consider and prioritise those who might be vulnerable to sustained unhealthy dietary changes (e.g. females, older individuals or the physically inactive), and those who face greater barriers to healthy change.

The new UK government obesity campaign needs to not only ensure that HFSS foods are less accessible (e.g. placing lower limits on cost or limiting advertising), but also ensure that healthier options such as fruit and vegetables are more accessible (i.e. cheaper or more readily available) [72,73]. Strategies need to consider the social impact of households on dietary behaviours and incorporating healthy eating into social norms and identities [74]. Accessible resources should be available for successful behavioural change and habit formation, promoting autonomy and satisfaction from healthful dietary changes [67].

### Strengths and limitations

There are several strengths of this study. This is one of the first studies in UK adults examining changes in dietary behaviours and predictors of dietary change across the first year of the COVID-19 pandemic, from May to December 2020 compared with pre-pandemic. The longitudinal nature builds upon the largely cross-sectional current literature, providing a greater understanding of the long-term impacts of the pandemic on dietary behaviour. The analysis included a range of variables that reflect the wide-ranging impact of the pandemic, with time-varying measures to reflect the changing conditions of the pandemic over time. A range of health behaviours were also considered that are important for dietary behaviour. The use of GEE models for the longitudinal analysis provided several advantages over common analytical methods, including the ability to handle repeated measures, model different data distributions and use time-varying predictors. Complete case analyses and sensitivity analyses with binary cut-offs demonstrating largely similar associations indicates the robustness of the associations identified in this study.

However, there are several limitations which may have introduced bias. Firstly, the study sample was self-selected and largely female, younger, and well-educated. Second, there were differences in various characteristics between included and excluded participants. This may limit the generalisability of results. Third, causality cannot be concluded from the observational study design. Fourth, measures of interest were self-reported; however epidemiological studies during COVID-19 have been largely self-reported, and dietary assessments in general are routinely self-reported [75]. The survey did not include a complete dietary analysis of all food groups or energy intake. Individuals generally tend to underestimate energy intake, and more so in individuals living with overweight or obesity [75,76]. However, this study focused on key food groups and their frequency of consumption, including HFSS foods and fruit and vegetables. Furthermore, self-reported dietary data still holds important value to inform health policy [77]. Using dietary change scores as the outcome variables enabled participants to act as their own controls which helped to minimise within-subject measurement error. Participants were also not explicitly asked if their diet had changed, nor told that dietary change was an outcome of interest, reducing the risk of expectation bias. Fifth, participants were asked about their behaviours in the past week or month, which may have introduced a recall bias. Sixth, the survey focused on HFSS snacking specifically, rather than any snacking. However, most studies to date have considered HFSS snacks [18].

## 5. Conclusion

While HFSS snacks intake fluctuated across the first year of the pandemic, it returned to pre-pandemic levels by the end of 2020. In contrast, HFSS meals also fluctuated but remained below pre-pandemic levels by November-December 2020. FV intake, while initially stable, decreased by the end of the year compared with pre-pandemic levels. These changes at population level do however, mask large interindividual changes in dietary behaviours, driven by differences in anthropometric (BMI), sociodemographic (gender), lifestyle (quality of life and living conditions) and behavioural (other dietary choices and physical activity levels) factors.

## Data Availability

Data are available upon request.

## Ethics

The study was approved by the Ethics Committee at the UCL Division of Psychology and Language Sciences (CEHP/2020/759). Participants gave their consent prior to data collection, and the study was GDPR compliant.

## Author contributions statement

S.D., J.J.M, J.N.L.V, E.B., D.K., A.H., L.S.; conceptualisation and methodology, E.B.; statistical support, S.D., J.J.M.; formal analysis, S.D.; first manuscript draft, L.S., D.K., A.H.; writing, reviewing and editing, A.H., L.S.; supervision. All authors approved the final manuscript.

## Informed Consent Statement

Informed consent was obtained from all subjects involved in the study.

