## Supplementary Materials for "Impact of COVID-19 pandemic on diet behaviour among UK adults: a longitudinal analysis of the HEBECO study"

### Substantive deviations from pre-registered protocol

The fruit and vegetable outcome variable for RQ2 was pre-registered as an ordinal, categorical change score (increased, no change, reduced intake) computed by comparing intakes at each timepoint (consuming a few portions per day vs all other) to pre-COVID-19 intakes retrospectively reported at baseline (consuming a few portions per day vs all other). However, upon formulating the outcome variable and running models, there was insufficient variance in the data, and GEE models were not converging. This was due to having no participants in the 'all other' ethnicity level for decreasing fruit and vegetable intake. In order to retain the change element of the fruit and vegetable outcome and maximise value for guiding health policy by identifying unhealthy dietary changes, the outcome variable for fruit and vegetable intake was changed to a binary, categorical change outcome (reduced intake vs all other) computed by comparing intakes at each timepoint (consuming a few portions per day vs all other) to pre-COVID-19 intakes retrospectively reported at baseline (consuming a few portions per day vs all other). The GEE models used the logit link function for a binomial distribution.

Given that dietary behaviours are likely to impact on other dietary behaviours, HFSS meals intake explanatory variable was included in the predictors for a change in HFSS snacks, and HFSS snacks was included in the predictors for a change in HFSS meals.

### Bayes Factors Robustness checks

Robustness checks were performed using alternative priors. For smoking status, a prior mean difference below -4.0 and above 11.0 HFSS snack portions per month produced the same outcome of no evidence for an effect.

### Testing linearity of logit in the binary sensitivity analysis models

Continuous explanatory variables were tested for linearity of logit, and converted to categorical variables where the assumption was violated.

For binary fruit and vegetables RQ2 and complete case analysis, physical activity violated the assumption.

- Physical activity: based on a lower limit cut-off of heath recommendations of 150 minutes per week of moderate or 75 minutes of vigorous physical activity equating to 7.5 MET-hours per week [1,2], as the minimum recommended activity levels. (Table 3, Supplementary tables S2, S4 and S5).

For HFSS snacks sensitivity increase or decrease vs all other, HFSS meals violated the assumption.

- HFSS meals median intake: below median vs median and above at each timepoint. Supplementary tables S6 and S7.

### Detailed description of study measures

The full wording of questions can be found at: https://osf.io/bja7g/

Time-invariant predictors are obtained from participants' baseline responses. Time-variant predictors are collected at baseline, 3- and 6-month follow-up surveys.

**Time-invariant**

**Socio-demographic**

- **Gender** (2 levels) *'Which of the following best describes how you think of yourself?'* **female** (female), **all others** (male, non-binary, prefer not to say)
- **Age.** continuous.
- **Ethnicity** (2 levels) **white** (1 white), **all others** (Mixed/Multiple ethnic groups, Asian/Asian British, Black/African/Caribbean/Black British, Chinese, Arab, Other ethnic group, prefer not to say)
- **Baseline occupation and working from home** (3 levels) **Unemployed, Employed from home, Employed not from home.**
  - All participants were asked, *'What is your current main occupation (during COVID-19)?'* Laid off during COVID-19, unemployed since before COVID-19, retired, homemaker, full-time parent or carer, unable to work due to disability, other, employed (full or part-time), self-employed (full or part-time), student, furloughed during COVID-19.
  - Participants who answered as employed (full or part-time), self-employed (full or part-time), student, furloughed during COVID-19 to the question *'What is your current main occupation (during COVID-19)?'* were asked, *'Can you do your work or study from home?'* Yes, I can do all the work or study from home, I can only do some work or study from home, or No, my work or study cannot be done at home.
  - **Unemployed** (Laid off during COVID-19, Unemployed since before COVID-19, Retired, Homemaker, Full-time parent or carer, Unable to work due to disability, Other), **Employed from home** (Employed AND Yes, I can do all the work or study from home, OR I can only do some work or study from home) **Employed not from home** (Employed AND No, my work or study cannot be done at home).
- **Composite socioeconomic score:** A sum score of education level and combined housing tenure and household income (Sum score 0-3):
  - **Education level.** *'What is the highest level of education that you have completed?'* **0,** below A-level (No formal qualification, GCSE/School certificate/O-level/CSE, Vocational qualifications (e.g. NVQ1+2)), or **1,** A-level or above (A-level/Higher school certificate or equivalent (e.g. NVQ3), Bachelor degree or equivalent (e.g. NVQ4), Masters/PhD/PGCE or equivalent, Other).
  - **Housing and Income.** *'What is your housing tenure?'* Owned outright, Mortgage Rented from local authority, Rented from private landlord, Belongs to housing association, Shared ownership (part owned, part rented), Other. '*What was your household annual income before COVID- 19?'* up to 13 499 GBP, 13 500-24 999 GBP, 25 000-49 999 GBP, ≥50 000 GBP, prefer not to say. **2** (Housing **=** Owned outright OR mortgage AND Income = 50 000+ GBP), **1** (Housing = All other AND Income = 50 000+ GBP), **1** (Housing = Owned outright OR mortgage AND Income = All other), **0** (Housing = All other AND Income = All other).

**COVID-19 related (impact on well-being and lifestyle)**

- **Living arrangements (3 levels): alone, with children (with or without adults), with adults only.**
  - **Alone.** Participants responding 'I live on my own' when asked *'How many persons other than yourself (including children) live with you now in the same flat or hous*e? Participants answering, 'I live with 1 or more people' were asked: *'Do you live with any of these persons below?'* Being able to select multiple responses: With my partner, husband/wife, boyfriend/girlfriend, with children 0-5 years old, with children 6-15 years old, with persons aged 16-69 (family or friends), with persons aged 70+ (family or friends), persons who you believe may be vulnerable to COVID-19 for any reason, persons who are in poor health

- **With any children (with or without adults)** (if participants respond with 'children 0-5 years old' or 'with children 6-15 years old')
- **Adults only** (all other participants who did not say 'I live on my own' or did not select 'with children 0-5 years old' or 'with children 6-15 years old')

**Time-variant**

**Socio-demographic**

- **BMI.** Continuous. Participants were asked at baseline, 3 months and 6 months later, *'How much do you weigh? Please try to be as accurate as possible'*. Participants could answer in 1lb/0.4-5kg increments, or answer 'don't know' or 'prefer not to say'. Participants were also asked *'What is your height? Please try to be as accurate as possible'*. Participants could answer in 1 inch/2-3 cm increments, or answer 'don't know' or 'prefer not to say'. BMI was calculated as weight (kg)/height$^2$ (m), by converting height into metres. Prefer not to say or don't know responses were excluded.

**COVID-19 related (impact on well-being and lifestyle)**

- **Isolation** *'Which type of COVID-19 induced isolation are you experiencing?'* (2 levels) **total and some** (Total isolation/quarantine, Some isolation), **general and no isolation** (General isolation, No isolation)
- **Composite wellbeing, social and living measure**: At baseline: *'How would you rate the below aspects of your life in the since COVID-19? (i) Living, (ii) wellbeing, (iii) social, (iv) family'.* At follow-up: *'How would you rate the below aspects of your life in the past month?'* 1-5: 1 poor, 2, 3 average, 4, 5 excellent. Continuous mean score of living + wellbeing + social + family (1-5)

**Behavioural**

- **Fruit and vegetables intake.** Continuous, from baseline-since-COVID-19 to 6 months. This predictor was used in the (i) HFSS snacks and (ii) HFSS meals intake analysis only.
- **HFSS snacks** and **HFSS meals intake.** Continuous, from baseline-since-COVID-19 to 6 months. These predictors were used in the (iii) fruit and vegetables intake analysis only.
- **Physical activity. Continuous.** A continuous measure of MET-hours per week from baseline-since-COVID-19 to 6 months was computed using weekly strength training and assuming a session length of 45 minutes, and using weekly aerobic training frequency and aerobic session length.
    - Weekly strength training. '*Since COVID-19, on average, on how many days per week have you done strength training?'* 0 days per week, 1 day per week, 2 days per week, 3 days per week, 4 days per week or more.
    - Weekly aerobic training. *'Since covid-19, how long (in minutes) was your average session of moderate or vigorous aerobic physical activity?'* (15-480 minutes) and '*Since covid-19 on average, how many times per week have you done 15 minutes or more of moderate or vigorous aerobic physical activity?'* (0,1,2 to 13 times per week, or 14+ times per week).
- **Alcohol consumption. Binary.** Participants were asked *'How often did you have a drink containing alcohol in the past month?', 'In the past month, how many units of alcohol did you drink on a typical day when you were drinking?'*. Scores were multiplied to compute a binary measure of weekly consumption, categorised into **≤14** and **>14** alcohol units.
- **Smoking.** At baseline, *'Which statement about tobacco use and cigarette smoking best describes you?'* At follow-ups, '*2 months ago/previously, we asked about your tobacco use. Your situation may have changed since then, or not. Which of the following best applies to you now?'* (2 levels) **Yes** (I smoke cigarettes (including hand-rolled) every day, I smoke cigarettes (including hand-rolled), but

not every day, I do not smoke cigarettes at all, but I do smoke tobacco of some kind (e.g. pipe, cigar or shisha), **No** (I have stopped smoking completely in the last year (at baseline)/in the past 2/3 months (follow-ups); I stopped smoking completely more than a year ago (baseline)/more than 2/3 months ago (follow-ups), I have never smoked any cigarettes/I have never been a regular smoker (i.e. smoked for a year or more).

**Supplementary tables and figures**

**Figure S1.** Changes in portion frequency consumption across the pandemic for (A) HFSS snacks intake, (B) HFSS meals intake and (C) fruit and vegetables intake.
**Figure S2.** Graphical illustrations of the time-varying associations of gender with meeting fruit and vegetable (FV) recommendations at baseline (May-June 2020), 3-months (August-September 2020) and 6-months follow-up (November-December 2020) from complete case analyses.
**Figure S3.** Graphical illustrations of the time-varying associations of (A) age and (B) alcohol consumption with increasing monthly HFSS snacks intake at baseline (May-June 2020), 3-months (August-September 2020) and 6-months follow-up (November-December 2020) from complete case analyses.
**Figure S4.** Graphical illustrations of the time-varying associations of eating behaviour score with a change in monthly (A) HFSS snacks and (B) meals intake at baseline (May-June 2020), 3-months (August-September 2020) and 6-months follow-up (November-December 2020) from complete case analyses.

**Table S1.** Weighted baseline participant characteristics for the total, included and excluded samples.
**Table S2.** Univariate unadjusted and adjusted models for each predictor variable and change in self-reported HFSS snacks and meals intake, and reduced FV intake.
**Table S3.** Unweighted mean changes in HFSS snacks and HFSS meals intake and changes in % meeting fruit and vegetable recommendations using complete cases.
**Table S4.** Univariate unadjusted and adjusted models for each predictor variable and change in self-reported HFSS snacks and HFSS meals intake, and reduced FV intake using complete cases.
**Table S5.** Fully adjusted GEE containing all predictor variables and the fully adjusted GEE model including significant explanatory variable*time interactions for a change in HFSS snacks and meals intake, and for reduced FV intake using complete cases.
**Table S6.** Univariate GEE models for a change in HFSS snacks intake using binary outcomes.
**Table S7.** Full GEE model containing all predictor variables and the full GEE model containing all predictor variables and significant explanatory variable*time interactions from univariate models adjusted for time, for a change in HFSS snacks intake with binary outcomes using complete cases.
**Table S8.** Univariate unadjusted and adjusted models for composite eating behaviour score and change in self-reported HFSS snacks and HFSS meals intake, and reduced FV intake using complete cases.
**Table S9.** Fully adjusted models for a change in HFSS snacks and meals intake, and for reduced FV intake using complete cases, including the eating behaviour score explanatory variable.

**Figure S1.** Changes in portion frequency consumption across the pandemic for (A) HFSS snacks intake, (B) HFSS meals intake and (C) fruit and vegetables intake.

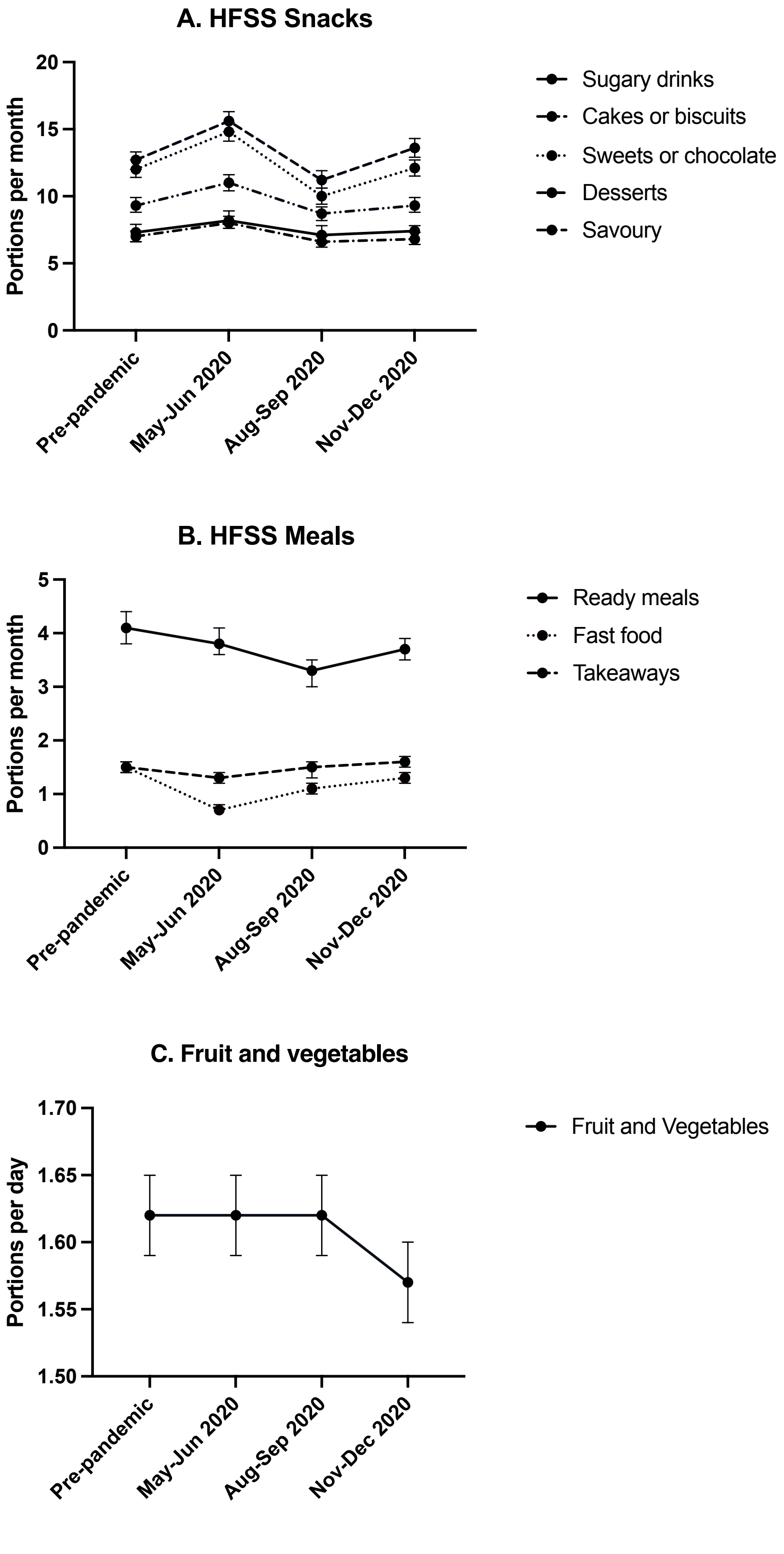

**Figure S2.** Graphical illustrations of the time-varying associations of gender with meeting fruit and vegetable recommendations at baseline (May-June 2020), 3-months (August-September 2020) and 6-months follow-up (November-December 2020) from complete case analyses.

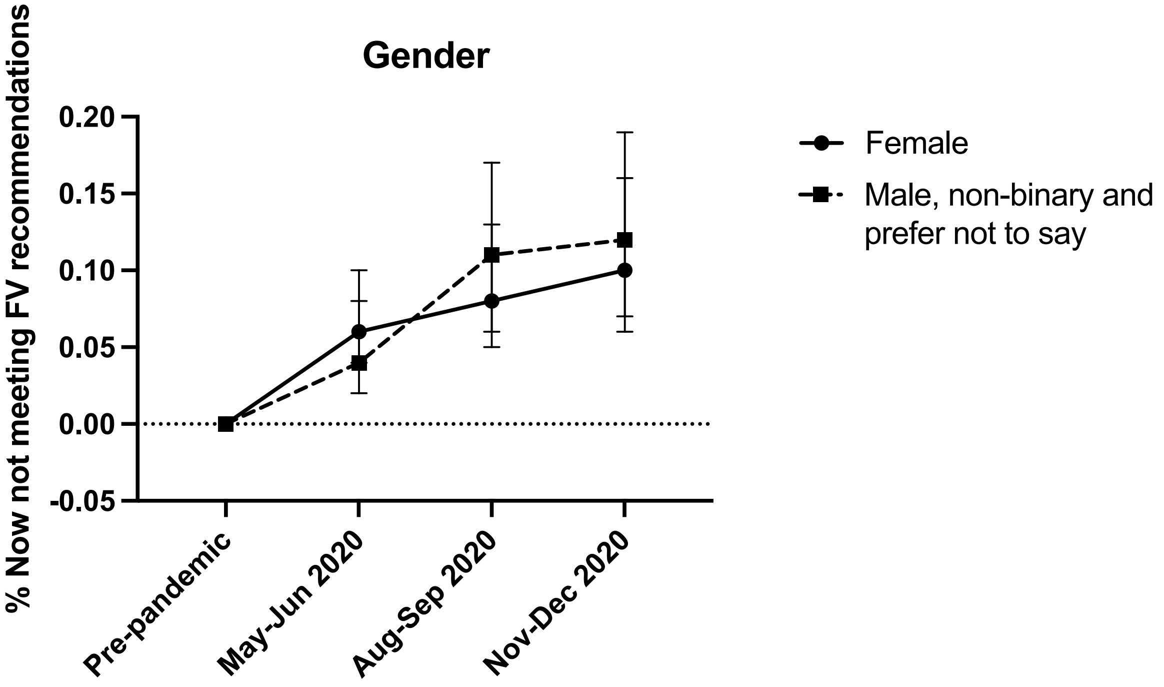

**Figure S3.** Graphical illustrations of the time-varying associations of (A) age and (B) alcohol consumption with increasing monthly HFSS snacks intake at baseline (May-June 2020), 3-months (August-September 2020) and 6-months follow-up (November-December 2020) from complete case analyses.

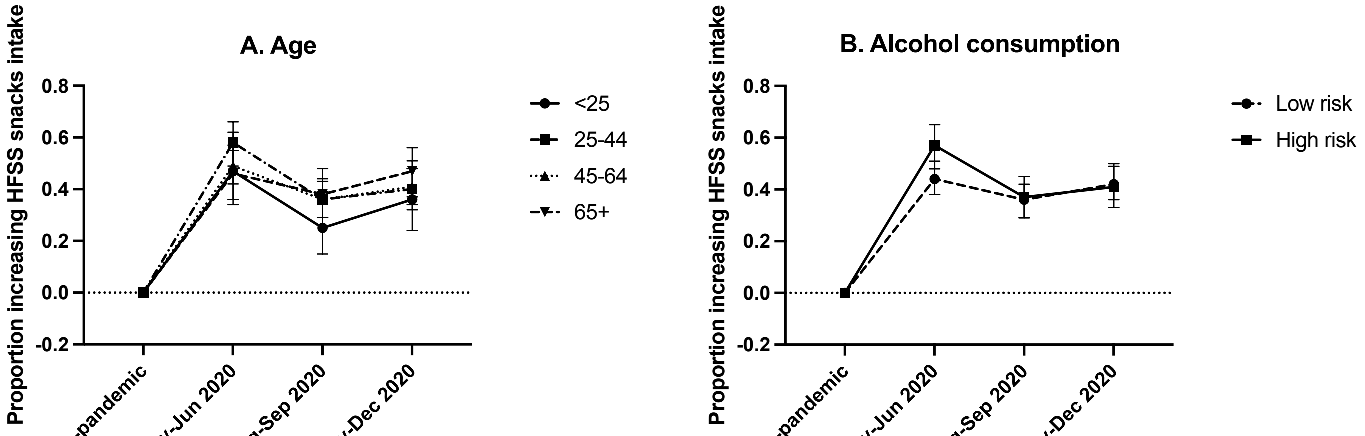

*Age categories: <25, 4.6%; 25-44, 23.1%; 45-64, 53.4%; >65, 19.0%.*

**Figure S4.** Graphical illustrations of the time-varying associations of eating behaviour score with a change in monthly (A) HFSS snacks and (B) meals intake at baseline (May-June 2020), 3-months (August-September 2020) and 6-months follow-up (November-December 2020) from complete case analyses.

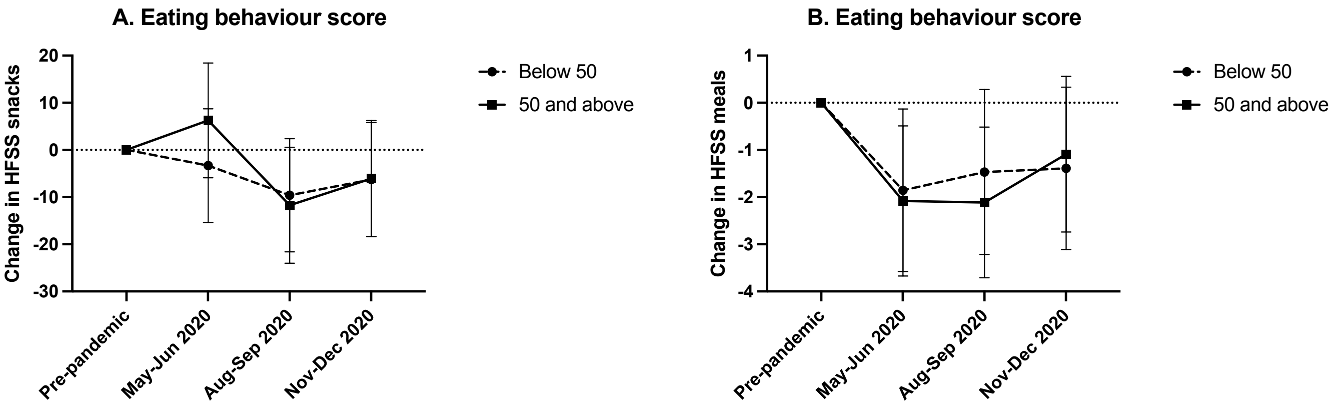

**Table S1.** Weighted baseline participant characteristics for the total, included and excluded samples.

| | Total Sample | Included sample | Excluded sample | |
|---|---|---|---|---|
| | Weighted (%) | Weighted (%) | Weighted (%) | p-value |
| N | 2805 | 1532 | 1274 | |
| Gender | | | | |
| All other | 48.3% | 45.8% | 51.3% | 0.004 |
| Female | 51.7% | 54.2% | 48.7% | |
| Ethnicity | | | | **<0.001** |
| All other | 11.5% | 8.0% | 15.7% | |
| White | 88.5% | 92.0% | 84.3% | |
| Mean BMI [SD] N= 2614 | 26.4 [5.2] | 26.7 [5.2] | 26.0 [5.2] | **<0.001** |
| Mean Age [SD] | 48.3 [16.7] | 52.9 [14.8] | 42.8 [17.2] | **<0.001** |
| Occupation and work from home N= 2806 | | | | **<0.001** |
| Unemployed (retired persons and full-time parents/carers) | 30.9% | 35.7% | 25.1% | |
| Employed working from home | 40.4% | 37.9% | 43.4% | |
| Employed not working from home | 28.7% | 26.4% | 31.5% | |
| Socioeconomic score | | | | **<0.001** |
| Income <£50k, unowned Housing and no higher education | 13.0% | 10.3% | 16.4% | |
| 1 of: ≥£50K income, housing ownership/mortgage or higher education | 40.5% | 39.2% | 42.0% | |
| 2 of: ≥£50K income, housing ownership/mortgage or higher education | 34.7% | 37.6% | 31.1% | |
| All of: ≥£50K income, housing ownership/mortgage and higher education | 11.8% | 12.9% | 10.5% | |
| Living conditions | | | | **<0.001** |
| Alone | 18.8% | 19.7% | 17.8% | |
| With children (with or without adults) | 17.3% | 14.4% | 20.7% | |
| With adults only | 63.9% | 65.9% | 61.5% | |
| Isolation status N= 2761 | | | | **<0.001** |
| Total or some isolation | 75.8% | 79.0% | 71.8% | |
| General or no isolation | 24.2% | 21.0% | 28.2% | |
| Mean Quality of Life [SD] N=2704 | 3.2 [0.9] | 3.3 [0.8] | 3.1 [0.9] | **<0.001** |
| Mean HFSS Snacks portions per month N=2395 | 61.1 [49.9] | 61.6 [50.5] | 60.3 [48.8] | 0.531 |
| Mean HFSS Meals portions per month N=2409 | 7.4 [9.2] | 6.6 [7.2] | 8.9 [11.8] | **<0.001** |
| Mean Fruit and vegetables portions per month N=2438 | 39.4 [19.7] | 40.5 [19.2] | 37.7 [20.3] | 0.001 |
| Mean physical activity MET-Hours per week N=2587 | 17.6 [19.8] | 18.3 [20.3] | 16.7 [19.1] | **0.040** |
| Alcohol intake N=2541 | | | | **0.044** |
| ≤14 weekly units | 80.8% | 82.1% | 78.9% | |
| >14 weekly units | 19.2% | 17.9% | 21.1% | |

| | | | | |
|---|---|---|---|---|
| Smoking status N=2806 | | | | **<0.001** |
| Yes | 26.5% | 18.6% | 35.9% | |
| No | 73.5% | 81.4% | 64.1% | |

*SD: Standard Deviation.*

**Table S2.** Univariate unadjusted and adjusted models for each predictor variable and change in self-reported HFSS snacks and meals intake, and reduced FV intake.

| | **Change in self-reported HFSS Snacks** | | | | **Change in self-reported HFSS meals** | | | | **Reduced FV intake** | | | |
|---|---|---|---|---|---|---|---|---|---|---|---|---|
| | **Unadjusted models** | | **Adjusted models w/interaction** | | **Unadjusted models** | | **Adjusted models w/interaction** | | **Unadjusted models** | | **Adjusted models w/interaction** | |
| **Predictor** | ***W χ2*** | **p** | ***W χ2*** | **p** | ***W χ2*** | **p** | ***W χ2*** | **p** | ***W χ2*** | **p** | ***W χ2*** | **p** |
| Gender | **12.220** | **<0.001** | **11.141** | **0.001** | **5.684** | **0.017** | **5.725** | **0.017** | 0.128 | 0.721 | 39.477 | 0.597 |
| Time | | | **192.667** | **<0.001** | | | **14.336** | **0.001** | | | **39.477** | **<0.001** |
| Time*Gender | | | 1.129 | 0.569 | | | 0.064 | 0.969 | | | **6.802** | **0.033** |
| Ethnicity | 0.544 | 0.461 | 0.697 | 0.404 | <0.001 | 0.974 | 0.001 | 0.974 | 0.004 | 0.950 | 0.017 | 0.897 |
| Time | | | **41.321** | **<0.001** | | | **7.316** | **0.026** | | | **10.01** | **0.007** |
| Time*Ethnicity | | | 1.585 | 0.453 | | | 1.032 | 0.597 | | | 0.790 | 0.674 |
| BMI | **5.278** | **0.022** | **5.152** | **0.023** | 1.282 | 0.258 | 1.820 | 0.177 | **15.118** | **<0.001** | **17.188** | **<0.001** |
| Time | | | **0.486** | **0.784** | | | 3.841 | 0.147 | | | **8.034** | **<0.001** |
| Time*BMI | | | **6.882** | **0.032** | | | 4.648 | 0.098 | | | 2.411 | 0.300 |
| Age | 0.001 | 0.976 | 0.293 | 0.588 | 0.005 | 0.945 | 0.020 | 0.886 | 2.264 | 0.132 | 2.319 | 0.128 |
| Time | | | **71.567** | **<0.001** | | | 5.089 | 0.079 | | | 2.271 | 0.321 |
| Time*Age | | | **17.175** | **<0.001** | | | 1.815 | 0.404 | | | 0.200 | 0.905 |
| Occupation and work from home | 0.788 | 0.674 | 0.224 | 0.894 | 0.357 | 0.836 | 0.217 | 0.897 | 1.684 | 0.431 | 2.586 | 0.274 |
| Time | | | **223.915** | **<0.001** | | | **14.285** | **0.001** | | | **35.595** | **<0.001** |
| Time*Occupation and work from home | | | 7.140 | 0.129 | | | 2.355 | 0.671 | | | 2.642 | 0.619 |

| | | | | | | | | | | | | |
|---|---|---|---|---|---|---|---|---|---|---|---|---|
| Socioeconomic score | 4.623 | 0.202 | 4.419 | 0.220 | 4.193 | 0.241 | **<0.001** | 1.000 | **9.647** | **0.022** | **8.712** | **0.033** |
| Time | | | **87.866** | **<0.001** | | | 10.964 | 0.004 | | | **20.027** | **<0.001** |
| Time*Socioeconomic score | | | **14.752** | **0.022** | | | 2.623 | 0.758 | | | 1.303 | 0.971 |
| Living arrangements | 1.084 | 0.582 | 0.962 | 0.618 | 5.822 | 0.054 | 5.991 | 0.050 | 3.505 | 0.173 | 2.838 | 0.242 |
| Time | | | **161.284** | **<0.001** | | | **21.977** | **<0.001** | | | **38.118** | **<0.001** |
| Time*Living arrangements | | | 6.349 | 0.175 | | | 4.819 | 0.306 | | | 4.057 | 0.398 |
| Isolation status | **81.509** | **<0.001** | 0.276 | 0.599 | 0.009 | 0.925 | 0.499 | 0.480 | **4.823** | **0.028** | **34.305** | **<0.001** |
| Time | | | **156.998** | **<0.001** | | | **22.932** | **<0.001** | | | **0.005** | **0.945** |
| Time*Isolation status | | | 1.528 | 0.466 | | | 0.346 | 0.841 | | | 1.942 | 0.379 |
| Quality of life | **39.870** | **<0.001** | **13.717** | **<0.001** | 0.005 | 0.946 | 0.193 | 0.661 | **9.897** | **0.002** | **12.070** | **0.001** |
| Time | | | **36.370** | **<0.001** | | | 4.75 | 0.093 | | | 1.453 | 0.484 |
| Time*Quality of life | | | **11.068** | **0.004** | | | 3.664 | 0.160 | | | 4.418 | 0.110 |
| HFSS snacks intake | | | | | **14.814** | **<0.001** | **16.668** | **<0.001** | 3.431 | 0.064 | **8.427** | **0.004** |
| Time | | | | | | | 0.329 | 0.848 | | | 25.778 | <0.0001 |
| Time*HFSS snacks intake | | | | | | | 3.969 | 0.137 | | | 2.215 | 0.330 |
| HFSS meals intake | **44.221** | **<0.001** | **48.416** | **<0.001** | | | | | **17.063** | **<0.001** | **6.591** | **0.010** |
| Time | | | **77.207** | **<0.001** | | | | | | | 30.703 | <0.001 |
| Time*HFSS meals intake | | | **18.409** | **<0.001** | | | | | | | 5.536 | 0.063 |

| | | | | | | | | | | | | |
|---|---|---|---|---|---|---|---|---|---|---|---|---|
| Fruit and vegetables intake | 0.165 | 0.685 | **<0.001** | 0.993 | 0.748 | 0.387 | 0.297 | 0.586 | | | | |
| Time | | | **66.725** | **<0.001** | | | **6.467** | **0.039** | | | | |
| Time*Fruit and vegetables intake | | | **15.214** | **<0.001** | | | 2.886 | 0.236 | | | | |
| Physical activity | **5.144** | **0.023** | **8.351** | **0.004** | 2.181 | 0.140 | 1.752 | 0.186 | **18.796** | **<0.001** | **15.154** | **<0.001** |
| Time | | | **157.374** | **<0.001** | | | **16.091** | **<0.001** | | | **41.646** | **<0.001** |
| Time*Physical activity | | | **8.200** | **0.017** | | | 2.896 | 0.235 | | | 0.097 | 0.953 |
| Alcohol intake | 0.001 | 0.970 | 1.790 | 0.181 | 0.138 | 0.710 | 0.114 | 0.735 | 0.242 | 0.622 | 0.079 | 0.779 |
| Time | | | **192.202** | **<0.001** | | | **15.872** | **<0.001** | | | **26.537** | **<0.001** |
| Time*Alcohol intake | | | 3.020 | 0.221 | | | 0.273 | 0.873 | | | 2.388 | 0.303 |
| Smoking status | 0.008 | 0.931 | 0.302 | 0.583 | 1.127 | 0.288 | 0.84 | 0.359 | 0.278 | 0.598 | 0.037 | 0.847 |
| Time | | | **117.28** | **<0.001** | | | **19.899** | **<0.001** | | | **29.553** | **<0.001** |
| Time*Smoking status | | | 2.352 | 0.309 | | | 5.043 | 0.080 | | | 1.934 | 0.380 |

*Missingness depends on if participants responded to survey questions relating to the predictor variable. For the binary fruit and vegetables analysis, physical activity violated the linearity of logit assumption, and was converted to a binary variable of below minimum health recommendations (<7.5 MET-hours per week) vs meeting minimum health recommendations (≥7.5 MET-hours per week). W $\chi 2$: Wald Chi-square.*

**Table S3.** Unweighted mean changes in HFSS snacks and HFSS meals intake and changes in the proportion meeting fruit and vegetable recommendations using complete cases.

| | N | Change in HFSS Snacks | | Change in HFSS Meals | | % Meeting FV recommendations | |
|---|---|---|---|---|---|---|---|
| | | **Mean difference [95% CI]** | **p** | **Mean difference [95% CI]** | **p** | **Mean difference % [95% CI]** | **p** |
| Pre-pandemic - May-Jun | 1486 | **9.2 [7.2,11.2]** | **<0.001** | **-1.3 [0.9,1.7]** | **<0.001** | 0.00 [-0.02,0.02] | 0.993 |
| Pre-pandemic - Aug-Sep | 1486 | **-4.9 [-6.8,-3.1]** | **<0.001** | **-1.3 [0.9,1.8]** | **<0.001** | 0.00 [-0.03,0.02] | 0.993 |
| Pre-pandemic - Nov-Dec | 1486 | 0.3 [-1.5,2.0] | 0.774 | **-0.7 [-1.2,-0.2]** | **0.001** | 0.02 [-0.01,0.05] | 0.379 |
| May-Jun - Aug-Sep | 1486 | **-14.1 [-16.4,-11.9]** | **<0.001** | -0.03 [-0.3,0.3] | 0.824 | 0.00 [-0.03,0.02] | 0.993 |
| Aug-Sep - Nov-Dec | 1486 | **5.2 [3.5,6.9]** | **<0.001** | **0.6 [0.2,1.0]** | **0.001** | 0.02 [-0.01,0.05] | 0.298 |
| May-Jun - Nov-Dec | 1486 | **-8.9 [-11.4,-6.4]** | **<0.001** | **0.6 [0.2,0.9]** | **0.001** | 0.02 [-0.01,0.05] | 0.379 |

*CI: Confidence Interval.*

**Table S4.** Univariate unadjusted and adjusted models for each predictor variable and change in self-reported HFSS snacks and HFSS meals intake, and reduced FV intake using complete cases.

| | Change in self-reported HFSS Snacks intake | | | | Change in self-reported HFSS meals intake | | | | Reduced FV intake | | | |
|---|---|---|---|---|---|---|---|---|---|---|---|---|
| | Unadjusted models | | Adjusted models w/interaction | | Unadjusted models | | Adjusted models w/interaction | | Unadjusted models | | Adjusted models w/interaction | |
| **Predictor** | ***W χ2*** | **p** | ***W χ2*** | **p** | ***W χ2*** | **p** | ***W χ2*** | **p** | ***W χ2*** | **p** | ***W χ2*** | **p** |
| Gender | **10.469** | **0.001** | **9.864** | **0.002** | **5.390** | **0.020** | **5.459** | **0.019** | 0.003 | 0.953 | 0.059 | 0.809 |
| Time | | | **187.119** | **<0.001** | | | **11.749** | **0.003** | | | **29.464** | **<0.001** |
| Time*Gender | | | 2.392 | 0.302 | | | 0.485 | 0.784 | | | **6.469** | **0.039** |
| Ethnicity | 0.975 | 0.323 | 1.147 | 0.284 | 2.930 | 0.087 | 3.211 | 0.073 | 0.001 | 0.976 | 0.073 | 0.787 |
| Time | | | 42.283 | **<0.001** | | | **8.162** | **0.017** | | | **8.293** | **0.016** |
| Time*Ethnicity | | | 1.125 | 0.570 | | | 1.332 | 0.514 | | | 1.293 | 0.524 |
| BMI | **4.141** | **0.042** | **4.475** | **0.034** | 1.443 | 0.230 | 1.839 | 0.175 | **19.17** | **<0.001** | **21.595** | **<0.001** |
| Time | | | 0.584 | 0.747 | | | 3.604 | 0.165 | | | 9.540 | 0.008 |
| Time*BMI | | | **6.827** | **0.033** | | | 4.717 | 0.095 | | | 4.259 | 0.119 |
| Age | 0.234 | 0.628 | 0.754 | 0.385 | 0.219 | 0.640 | 0.411 | 0.522 | 0.596 | 0.440 | 0.605 | 0.437 |
| Time | | | **68.023** | **<0.001** | | | 3.157 | 0.206 | | | 2.334 | 0.311 |
| Time*Age | | | **16.053** | **<0.001** | | | 1.533 | 0.465 | | | 0.007 | 0.997 |
| Occupation and work from home | 0.156 | 0.925 | 0.080 | 0.961 | 0.973 | 0.615 | 0.682 | 0.711 | 1.337 | 0.512 | 1.631 | 0.442 |
| Time | | | **218.606** | **<0.001** | | | **9.808** | **0.007** | | | **24.722** | **<0.001** |
| Time*Occupation and work from home | | | **9.701** | **0.046** | | | 3.857 | 0.426 | | | 1.426 | 0.840 |

| | | | | | | | | | | | | |
|---|---|---|---|---|---|---|---|---|---|---|---|---|
| Socioeconomic score | 2.476 | 0.480 | ** | ** | 2.216 | 0.529 | 1.977 | 0.577 | **6.075** | **0.108** | ** | ** |
| Time | | | **73.356** | **<0.001** | | | **9.516** | **0.009** | | | ** | ** |
| Time*Socioeconomic score | | | 7.292 | 0.063 | | | 5.578 | 0.472 | | | ** | ** |
| Living arrangements | 1.066 | 0.587 | 1.058 | 0.589 | 5.101 | 0.078 | 5.339 | 0.069 | 5.408 | 0.067 | 4.758 | 0.093 |
| Time | | | **154.476** | **<0.001** | | | **12.034** | **0.002** | | | **26.309** | **<0.001** |
| Time*Living arrangements | | | 8.838 | 0.065 | | | 2.132 | 0.711 | | | 4.47 | 0.346 |
| Isolation status | **94.128** | **<0.001** | 0.131 | 0.718 | 0.058 | 0.810 | 0.590 | 0.442 | 2.326 | 0.127 | 0.313 | 0.576 |
| Time | | | **144.098** | **<0.001** | | | **16.252** | **<0.001** | | | **23.867** | **<0.001** |
| Time*Isolation status | | | 1.526 | 0.466 | | | 0.154 | 0.926 | | | 1.987 | 0.370 |
| Quality of life | **37.700** | **<0.001** | **11.477** | **0.001** | 0.030 | 0.862 | 0.131 | 0.717 | **6.844** | **0.009** | **8.791** | **0.003** |
| Time | | | **34.275** | **<0.001** | | | 4.228 | 0.121 | | | 1.183 | 0.553 |
| Time*Quality of life | | | **10.814** | **0.004** | | | 3.745 | 0.154 | | | 2.718 | 0.257 |
| HFSS snacks intake | | | | | **16.672** | **<0.001** | **18.789** | **<0.001** | 3.414 | 0.065 | **7.774** | **0.005** |
| Time | | | | | | | 0.766 | 0.682 | | | **16.548** | **<0.001** |
| Time*HFSS snacks intake | | | | | | | 1.857 | 0.395 | | | 1.419 | 0.492 |
| HFSS meals intake | **27.342** | **<0.001** | **35.700** | **<0.001** | | | | | 14.570 | **<0.001** | **7.007** | **0.008** |
| Time | | | **65.614** | **<0.001** | | | | | | | **19.123** | **<0.001** |
| Time*HFSS meals intake | | | **18..077** | **<0.001** | | | | | | | **6.700** | **0.035** |

| | | | | | | | | | | | | |
|---|---|---|---|---|---|---|---|---|---|---|---|---|
| Fruit and vegetables intake | 0.054 | 0.816 | 0.222 | 0.638 | 0.510 | 0.475 | 0.294 | 0.588 | | | | |
| Time | | | **61.166** | **<0.001** | | | 4.772 | 0.092 | | | | |
| Time*Fruit and vegetables intake | | | **13.087** | **0.001** | | | 2.355 | 0.308 | | | | |
| Physical activity (MET-hours per week) | **5.039** | **0.025** | **9.338** | **0.002** | 2.598 | 0.107 | 2.310 | 0.129 | **19.970** | **<0.001** | **15.338** | **<0.001** |
| Time | | | **148.862** | **<0.001** | | | **7.461** | **0.024** | | | **28.332** | **<0.001** |
| Time*Physical activity | | | **8.271** | **0.016** | | | 0.323 | 0.851 | | | 1.390 | 0.499 |
| Alcohol consumption | 0.159 | 0.690 | 0.688 | 0.407 | 0.008 | 0.929 | 0.005 | 0.945 | 0.001 | 0.973 | 0.002 | 0.966 |
| Time | | | **182.269** | **<0.001** | | | **15.732** | **<0.001** | | | **16.573** | **<0.001** |
| Time*Alcohol consumption | | | 4.165 | 0.125 | | | 0.559 | 0.756 | | | 1.790 | 0.409 |
| Smoking status | 0.614 | 0.433 | 1.267 | 0.260 | 2.307 | 0.129 | 1.867 | 0.172 | **1.515** | **0.218** | **0.744** | **0.388** |
| Time | | | **113.324** | **<0.001** | | | **9.506** | **0.009** | | | **21.952** | **<0.001** |
| Time*Smoking status | | | 4.617 | 0.099 | | | 1.165 | 0.559 | | | 3.022 | 0.221 |

***Model did not compute due to numerical errors. For the binary fruit and vegetables analysis, physical activity violated the linearity of logit assumption, and was converted to a binary variable of below minimum health recommendations (<7.5 MET-hours per week) vs meeting minimum health recommendations (≥7.5 MET-hours per week). Missingness with each model depends on if participants responded to survey questions relating to the predictor variable. W $\chi 2$: Wald Chi-square.*

**Table S5.** Fully adjusted GEE containing all predictor variables and the fully adjusted GEE model including significant predictor*time interactions for a change in HFSS snacks and meals intake, and for reduced FV intake using complete cases.

| | **Change in self-reported HFSS Snacks QIC = 4214122.36** | | **Change in self-reported HFSS Meals QIC = 184695.497** | | **Reduced FV intake QIC = 2282.216** | |
|---|---|---|---|---|---|---|
| All predictors (N=1451) | **p-value** | **B [95% CI]** | **p-value** | **B [95% CI]** | **p-value** | **OR [95% CI]** |
| Gender | **<0.001** | | **0.010** | | 0.737 | |
| All other | | **Reference** | | **Reference** | | Reference |
| Female | | **6.210 [3.127,9.294]** | | **0.948 [0.227,1.668]** | | 0.949 [0.697,1.290] |
| Ethnicity | 0.110 | | 0.064 | | 0.753 | |
| All other | | Reference | | Reference | | Reference |
| White | | -6.051 [-13.483,1.380] | | -1.123 [-2.312,0.065] | | 0.889 [0.426,1.853] |
| BMI | 0.166 | 0.233 [-0.097,0.564] | 0.084 | -0.056 [-0.119,0.008] | **0.006** | **1.035 [1.010,1.061]** |
| Age | 0.062 | 0.126 [-0.006,0.259] | 0.496 | 0.010 [-0.018,0.038] | 0.625 | 0.997 [0.984,1.010] |
| Occupation and work from home | 0.597 | | 0.692 | | 0.655 | |
| Unemployed | | Reference | | Reference | | Reference |
| Employed working from home | | 1.972 [-1.894,5.839] | | 0.077 [-0.982,0.828] | | 1.204 [0.811,1.788] |
| Employed not working from home | | 1.626 [-3.080,6.332] | | -0.412 [-1.485,0.661] | | 1.145 [0.727,1.805] |
| Socioeconomic score | 0.258 | | 0.384 | | 0.410 | |
| Income <£50k, unowned Housing and no higher education | | Reference | | Reference | | Reference |
| 1 of: ≥£50K income, housing ownership/mortgage or higher education | | 0.684 [-10.392,11.761] | | 1.739 [-1.103,4.580] | | 1.238 [0.473,3.240] |
| 2 of: ≥£50K income, housing ownership/mortgage or higher education | | 4.401 [-6.592,15.393] | | 2.076 [-0.695,4.846] | | 1.420 [0.550,3.665] |
| All of: ≥£50K income, housing ownership/mortgage and higher education | | 3.692 [-7.167,14.551] | | 2.134 [-0.629,4.897] | | 1.078 [0.408,2.852] |
| Living conditions | 0.473 | | **0.039** | | 0.938 | |
| Alone | | Reference | | **Reference** | | Reference |
| With children (with or without adults) | | 2.181 [-3.082,7.444] | | **-0.476 [-1.901,0.948]** | | 0.990 [0.586,1.672] |

| | | | | | | |
|---|---|---|---|---|---|---|
| With adults only | | 2.439 [-1.471,6.348] | | **0.658 [-0.375,1.691]** | | 0.943 [0.640,1.390] |
| Isolation status | 0.507 | | 0.373 | | 0.709 | |
| Total or some isolation | | Reference | | Reference | | Reference |
| General or no isolation | | -0.711[-2.813,1.391] | | 0.188 [-0.226,0.602] | | 1.053 [0.803,1.380] |
| Quality of Life | **<0.001** | **-2.785 [-4.318,-1.252]** | 0.971 | 0.005 [-0.288,0.299] | 0.098 | 0.867 [0.731,1.027] |
| HFSS Snacks | | | **<0.001** | **0.018 [0.010,0.027]** | 0.324 | 1.002 [0.998,1.005] |
| HFSS Meals | **<0.001** | **0.731 [0.453,1.008]** | | | 0.142 | 1.013 [0.996,1.031] |
| Fruit and vegetables | 0.401 | 0.034 [-0.045,0.112] | 0.306 | -0.008 [-0.025,0.008] | | |
| Physical activity (MET-Hours per week) | 0.102 | -0.036 [-0.080,0.007] | 0.074 | -0.009 [-0.018,0.001] | **0.004** | **0.671 [0.511,0.881]*** |
| Alcohol intake | 0.141 | | 0.441 | | 0.960 | |
| ≤14 weekly units | | Reference | | Reference | | Reference |
| >14 weekly units | | 1.659 [-0.548,3.867] | | 0.186 [-0.287,0.658] | | 1.008 [0.740,1.373] |
| Smoking status | 0.238 | | 0.283 | | 0.544 | |
| Yes | | Reference | | Reference | | Reference |
| No | | 2.600 [-1.720,6.920] | | -0.461 [-1.304,0.381] | | 0.884 [0.592,1.318] |
| **All predictors + significant full model interactions** | **Change in self-reported HFSS Snacks QIC = 4187090.57** | | | | **Reduced FV intake QIC = 2280.087** | |
| | **p-value** | | | | **p-value** | |
| Time*Age | **0.016** | | | | | |
| Time*HFSS meals intake | **0.001** | | | | | |
| Time*Gender | | | | | **0.042** | |

*For the fully adjusted GEE model including explanatory variable*time interactions, Type III tests for the explanatory variable*time interactions are shown only. QIC is a relative, 'lower is better' measure of goodness of fit. For the binary fruit and vegetables analysis, physical activity violated the linearity of logit assumption, and was converted to a binary variable of below minimum health recommendations (<7.5 MET-hours per week) vs meeting minimum health recommendations (≥7.5 MET-hours per week). *OR for 'meeting minimum health recommendations' with 'below minimum health recommendations' as reference. W χ2: Wald Chi-square, $B$: Beta parameter, SE: Standard Error, CI: Confidence Interval, OR: Odds Ratio.*

**Table S6.** Univariate GEE models for a change in HFSS snacks intake using binary outcomes.

| | Change in self-reported HFSS Snacks | | | | | | | |
|---|---|---|---|---|---|---|---|---|
| | Increase vs all other | | | | Decrease vs all other | | | |
| | Unadjusted models | | Adjusted models w/interaction | | Unadjusted models | | Adjusted models w/interaction | |
| **Predictor** | ***W χ2*** | **p-value** | ***W χ2*** | **p-value** | ***W χ2*** | **p-value** | ***W χ2*** | **p-value** |
| Gender | **11.269** | **<0.001** | **10.735** | **0.001** | **8.883** | **0.003** | **9.027** | **0.003** |
| Time | | | **53.031** | **<0.001** | | | **190.436** | **<0.001** |
| Time*Gender | | | 3.305 | 0.192 | | | 2.311 | 0.315 |
| Ethnicity | 0.004 | 0.949 | 0.003 | 0.956 | 0.054 | 0.816 | 0.072 | 0.788 |
| Time | | | **6.970** | **0.031** | | | **31.307** | **<0.001** |
| Time*Ethnicity | | | 1.272 | 0.529 | | | 0.839 | 0.657 |
| BMI | **4.782** | **0.029** | **4.851** | **0.028** | 0.348 | 0.555 | 0.640 | 0.424 |
| Time | | | 0.360 | 0.835 | | | 6.840 | 0.033 |
| Time*BMI | | | 1.332 | 0.514 | | | 0.082 | 0.960 |
| Age | 1.107 | 0.293 | 0.662 | 0.416 | 2.363 | 0.124 | 2.572 | 0.109 |
| Time | | | **30.120** | **<0.001** | | | **25.181** | **<0.001** |
| Time*Age | | | **15.183** | **0.001** | | | 1.816 | 0.403 |
| Occupation and work from home | 1.139 | 0.566 | 0.822 | 0.663 | 0.675 | 0.713 | 0.827 | 0.661 |
| Time | | | **56.224** | **<0.001** | | | **207.576** | **<0.001** |
| Time*Occupation and work from home | | | 8.393 | 0.078 | | | 5.067 | 0.280 |

| | | | | | | | | |
|---|---|---|---|---|---|---|---|---|
| Socioeconomic score | 3.288 | 0.349 | 2.850 | 0.415 | **8.228** | **0.042** | **8.367** | **0.039** |
| Time | | | **23.238** | **<0.001** | | | **92.627** | **<0.001** |
| Time*Socioeconomic score | | | 8.147 | 0.228 | | | 1.341 | 0.969 |
| Living arrangements | 1.864 | 0.394 | 2.180 | 0.336 | **6.675** | **0.036** | 7.481 | **0.024** |
| Time | | | **47.547** | **<0.001** | | | **150.283** | **<0.001** |
| Time*Living arrangements | | | 7.303 | 0.121 | | | 6.351 | 0.174 |
| Isolation status | **34.294** | **<0.001** | 1.266 | 0.261 | **74.726** | **<0.001** | 0.104 | 0.747 |
| Time | | | **31.881** | **<0.001** | | | **141.312** | **<0.001** |
| Time*Isolation status | | | 3.523 | 0.172 | | | 0.396 | 0.821 |
| Quality of life | **15.749** | **<0.001** | **6.046** | **0.014** | **13.323** | **<0.001** | 2.132 | 0.144 |
| Time | | | **8.893** | **0.012** | | | **11.559** | **0.003** |
| Time*Quality of life | | | 2.466 | 0.291 | | | 0.098 | 0.952 |
| HFSS meals intake | **19.176** | **<0.001** | **23.181** | **<0.001** | **16.208** | **<0.001** | **24.190** | **<0.001** |
| Time | | | **73.444** | **<0.001** | | | **240.892** | **<0.001** |
| Time*HFSS meals intake | | | 2.692 | 0.260 | | | 2.901 | 0.234 |
| Fruit and vegetables | 0.195 | 0.658 | 0.079 | 0.779 | 0.511 | 0.475 | 0.742 | 0.389 |
| Time | | | **12.075** | **0.002** | | | **34.272** | **<0.001** |
| Time*Fruit and vegetables | | | 0.576 | 0.750 | | | 1.061 | 0.588 |

| | | | | | | | | |
|---|---|---|---|---|---|---|---|---|
| Physical activity | 1.577 | 0.209 | 2.153 | 0.142 | 0.019 | 0.891 | 0.500 | 0.480 |
| Time | | | **34.026** | **<0.001** | | | **113.971** | **<0.001** |
| Time*Physical activity | | | 1.658 | 0.437 | | | 0.011 | 0.994 |
| Alcohol consumption | 0.298 | 0.585 | 1.182 | 0.277 | 0.182 | 0.670 | **3.340** | **0.068** |
| Time | | | **63.287** | **<0.001** | | | **156.775** | **<0.001** |
| Time*Alcohol consumption | | | **8.884** | **0.012** | | | 5.309 | 0.070 |
| Smoking status | 0.261 | 0.609 | 0.096 | 0.756 | 0.049 | 0.825 | 0.085 | 0.770 |
| Time | | | **33.454** | **<0.001** | | | **112.573** | **<0.001** |
| Time*Smoking status | | | 2.868 | 0.238 | | | 3.099 | 0.212 |

*Due to violating the linearity of logit assumption, HFSS meals intake was converted to a binary variable of 'below median' and 'median and above' intakes. Missingness with each model depends on if participants responded to survey questions relating to the predictor variable. W $\chi 2$: Wald Chi-square.*

**Table S7.** Full GEE model containing all predictor variables and the full GEE model containing all predictor variables and significant explanatory variable*time interactions from univariate models adjusted for time, for a change in HFSS snacks intake using binary outcomes using complete cases.

| | Change in self-reported HFSS snacks intake | | | | | |
|---|---|---|---|---|---|---|
| | Increase vs all other | | | Decrease vs all other | | |
| | Type III Test | | | Type III Test | | |
| **N=1451** | ***W χ2*** | **p-value** | **QIC** | ***W χ2*** | **p-value** | **QIC** |
| All predictors | | | | | | |
| | | | 5588.258 | | | 5278.921 |
| Gender | **15.120** | **<0.001** | | **12.365** | **<0.001** | |
| Ethnicity | 0.427 | 0.514 | | 1.419 | 0.234 | |
| BMI | **4.057** | **0.044** | | 0.028 | 0.867 | |
| Age | 0.014 | 0.905 | | **10.347** | **0.001** | |
| Occupation and work from home | 0.753 | 0.686 | | 2.768 | 0.251 | |
| Socioeconomic score | 4.697 | 0.195 | | 6.537 | 0.088 | |
| Living conditions | 1.049 | 0.592 | | 5.08 | 0.079 | |
| Isolation status | 1.648 | 0.199 | | 0.704 | 0.401 | |
| Quality of Life | **7.634** | **0.006** | | **7.022** | **0.008** | |
| HFSS meals intake | **19.344** | **<0.001** | | **24.292** | **<0.001** | |
| Fruit and vegetables | 0.003 | 0.958 | | 0.837 | 0.360 | |
| Physical activity (MET-Hours per week) | 0.073 | 0.788 | | 0.066 | 0.798 | |
| Alcohol consumption | 3.161 | 0.075 | | **5.442** | **0.020** | |
| Smoking status | 0.001 | 0.970 | | 0.017 | 0.896 | |
| **All predictors and significant interactions: N=1451** | | | 5574.692 | | | |
| Time*Age | 14.137 | **0.001** | | | | |
| Time*Alcohol consumption | 9.825 | **0.007** | | | | |

*Models also included Time as a covariate. Type III tests are shown only. Due to violating the linearity of logit assumption, HFSS meals intake was converted to a binary variable of 'below median' and 'median and above' intakes. There were no material changes in significance of main effects with the addition of time interactions, except for baseline BMI for an increase in HFSS snacks vs all other, which became non-significant. QIC is a relative, 'lower is better' measure of goodness of fit. W χ2: Wald Chi-square.*

**Table S8.** Univariate unadjusted and adjusted models for composite eating behaviour score and change in self-reported HFSS snacks and HFSS meals intake, and reduced FV intake using complete cases.

| | **Change in self-reported HFSS Snacks intake** | | | | **Change in self-reported HFSS meals intake** | | | | **Reduced FV intake** | | | |
|---|---|---|---|---|---|---|---|---|---|---|---|---|
| | **Unadjusted models** | | **Adjusted models w/interaction** | | **Unadjusted models** | | **Adjusted models w/interaction** | | **Unadjusted models** | | **Adjusted models w/interaction** | |
| **Predictor** | **W X-S** | **p** | **W X-S** | **p** | **W X-S** | **p** | **W X-S** | **p** | **W X-S** | **p** | **W X-S** | **p** |
| Composite eating behaviour score N=1193 | **25.474** | **<0.001** | **18.645** | **<0.001** | 0.204 | 0.652 | <0.001 | 0.984 | **6.099** | **0.014** | **6.248** | **0.012** |
| Time | | | **10.456** | **0.005** | | | **14.720** | **0.001** | | | 5.914 | 0.052 |
| Time*Composite eating behaviour score | | | **72.869** | **<0.001** | | | **9.258** | **0.010** | | | 0.246 | 0.884 |

*W χ2: Wald Chi-square.*

**Table S9.** Fully adjusted models for a change in HFSS snacks and meals intake, and for reduced FV intake using complete cases, including eating behaviour score.

| | **Change in self-reported HFSS Snacks** | | **Change in self-reported HFSS Meals** | | **Reduced FV intake** | |
|---|---|---|---|---|---|---|
| | **QIC = 3183339.18** | | **QIC = 135661.533** | | **QIC = 1802.887** | |
| All predictors (N=1166) | **W $\chi 2$** | **p-value** | **W $\chi 2$** | **p-value** | **W $\chi 2$** | **p-value** |
| Eating Behaviour score | **13.558** | **<0.001** | 0.118 | 0.731 | 0.326 | 0.568 |
| All predictors and significant interactions (N=1166) | **Change in self-reported HFSS Snacks** | | **Change in self-reported HFSS Meals** | | | |
| | **QIC = 3127023.43** | | **QIC = 135368.316** | | | |
| Time*Eating Behaviour score | **50.783** | **<0.001** | **7.351** | **0.025** | | |

*Models also included Time as a covariate. Type III tests are shown only for Eating behaviour score in the model. QIC is a relative, 'lower is better' measure of goodness of fit. W $\chi 2$: Wald Chi-square.*

[2] Kaminsky LA, Montoye AHK. Physical Activity and Health: What Is the Best Dose? J Am Heart Assoc n.d.;3:e001430. https://doi.org/10.1161/JAHA.114.001430.
